# Genotyping and population structure of the China Kadoorie Biobank

**DOI:** 10.1101/2022.05.02.22274487

**Authors:** Robin G Walters, Iona Y Millwood, Kuang Lin, Dan Schmidt Valle, Pandora McDonnell, Alex Hacker, Daniel Avery, Na Cai, Warren W Kretzschmar, M Azim Ansari, Paul A Lyons, Rory Collins, Peter Donnelly, Michael Hill, Richard Peto, Hongbing Shen, Xin Jin, Chao Nie, Xun Xu, Yu Guo, Canqing Yu, Jun Lv, Robert J Clarke, Liming Li, Zhengming Chen, the China Kadoorie Biobank Collaborative Group

## Abstract

China Kadoorie Biobank is a population-based prospective cohort of >512,000 adults recruited in 2004-2008 from 10 geographically diverse regions across China. Detailed data from questionnaire and physical measurements were collected at baseline, with additional measurements at three resurveys involving approximately 5% of surviving participants. Incident disease events are captured through electronic linkage to death and disease registries and to the national health insurance system. Genome-wide genotyping has been conducted for >100,000 participants using custom-designed Axiom^®^ arrays. Analysis of these data reveals extensive relatedness within the CKB cohort, signatures of recent consanguinity, and principal component signatures reflecting large-scale population movements from recent Chinese history. In addition to numerous CKB studies of candidate drug targets and disease risk factors, CKB has made substantial contributions to many international genetics consortia. Collected biosamples are now being used for high-throughput ‘omics assays which, together with planned whole genome sequencing, will continue to enhance the scientific value of this biobank.

## Introduction

Major non-communicable chronic diseases, such as heart attack, stroke, cancer, and chronic obstructive pulmonary disease (COPD), account for much of the adult disease burden in China and globally. Several such diseases display large unexplained variations in incidence between different regions in China, indicating that important genetic and non-genetic causes remain to be discovered. The China Kadoorie Biobank (CKB) was initiated in 2002, with the goal of investigating the causal relevance of established and novel disease risk factors in the adult Chinese population^1^. During 2004-2008, CKB recruited >512,000 adults aged 30-79 years from 10 geographically-diverse (five urban, five rural) regions across China, making it one of the largest blood-based prospective biobanks in the world^2^.

Many aspects of the CKB study design support a wide range of hypothesis-driven and hypothesis-free research into many different diseases: population-based recruitment; prospective sample collection; a relatively medication-naïve population; rich and diverse exposure and lifestyle data; and comprehensive capture of incident disease events through electronic linkage to death and disease registries and to health insurance records. CKB also contributes to the growing demand for ancestrally-diverse biobanks, which not only expand opportunities for novel discoveries of potential value to all human populations but also address potential inequalities in healthcare that may arise from the historical focus of research on individuals of European ancestry, findings from which are not necessarily transferable to other populations^3,4^. In common with many other large biobanks, the value of CKB has been greatly enhanced by large-scale genotyping of study participants. Such genotype information enables investigation of the contribution of genetic variation to phenotype and disease risk, Mendelian randomisation (MR) assessment of the causal contribution of risk factors and behaviours to disease, and phenome-wide analyses of the impact of variation at specific loci.

We describe the design and performance of a custom Affymetrix Axiom^®^ array optimised for individuals of Chinese Han ancestry, which provides both genome-wide coverage to enable high-quality imputation of both common and low-frequency variation, and direct genotyping of ∼68,000 putative loss-of-function, missense, and eQTL variants of potential use for MR or phenome-wide association studies. Based on genotyping of >100,000 CKB participants, we demonstrate extensive population diversity across China, identify substantial relatedness within the CKB study population, and observe principal component (PC) signatures consistent with population movements from recent Chinese history. Through linkage to deep phenotyping and >1.2M recorded disease events in CKB, this genotyping has already enabled a wide range of studies, from investigation of ancestry-specific loss of function variants to inform drug target identification, validation, and repurposing^5-8^; to participation in international trans-ancestry genome-wide association study (GWAS) consortia including the Global Biobank Meta-analysis Initiative (GBMI)^9-14^.

### Study Population and Data Collection

Recruitment of CKB participants was community-based, taking place at a large number of local ‘clinics’ within each of 10 diverse regions of China. At baseline assessment participants completed an extensive interviewer-administered questionnaire on factors including demographics and socio-economic status, diet and lifestyle (e.g. smoking, alcohol), physical activity, reproductive history (for women), and medical history and current medication. In addition, all participants underwent physical examination including measurements of anthropometrics, blood pressure, spirometry, exhaled carbon monoxide, and body composition (using bioimpedance). Furthermore, there were onsite blood tests of (non-fasting) glucose and hepatitis B surface antigen, and blood samples were processed within a few hours to separate plasma and buffy coat for long-term storage^2^.

Subsequent to initial recruitment, three periodic resurveys have been undertaken of approximately 5% of surviving participants, selected on the basis of representative random samples of recruitment clinics, to provide repeat measurements for correction of regression dilution bias, gather additional questionnaire information, conduct additional physical measurements and blood tests, and collect repeat blood samples and other additional biosamples for long-term storage (**Figure 1**). The first resurvey, conducted immediately after completion of study recruitment in 2008, was largely a repeat of the baseline survey^2^. This was extended in the second resurvey, conducted in 2013-2014, with additional questionnaire data, additional physical measurements, on-site assays of blood lipids, and collection, testing, and storage of urine samples. Further extensions in the third resurvey (2020-2021), which was mostly conducted in the same locations as the second, included additional measurements of abdominal ultrasound and retinal imaging, and collection of saliva and stool samples. Over 15,000 individuals attended at least two resurveys; these multiple measurements at different time points will enable future longitudinal analyses of trajectories of risk factors for major diseases.

**Figure 1.**
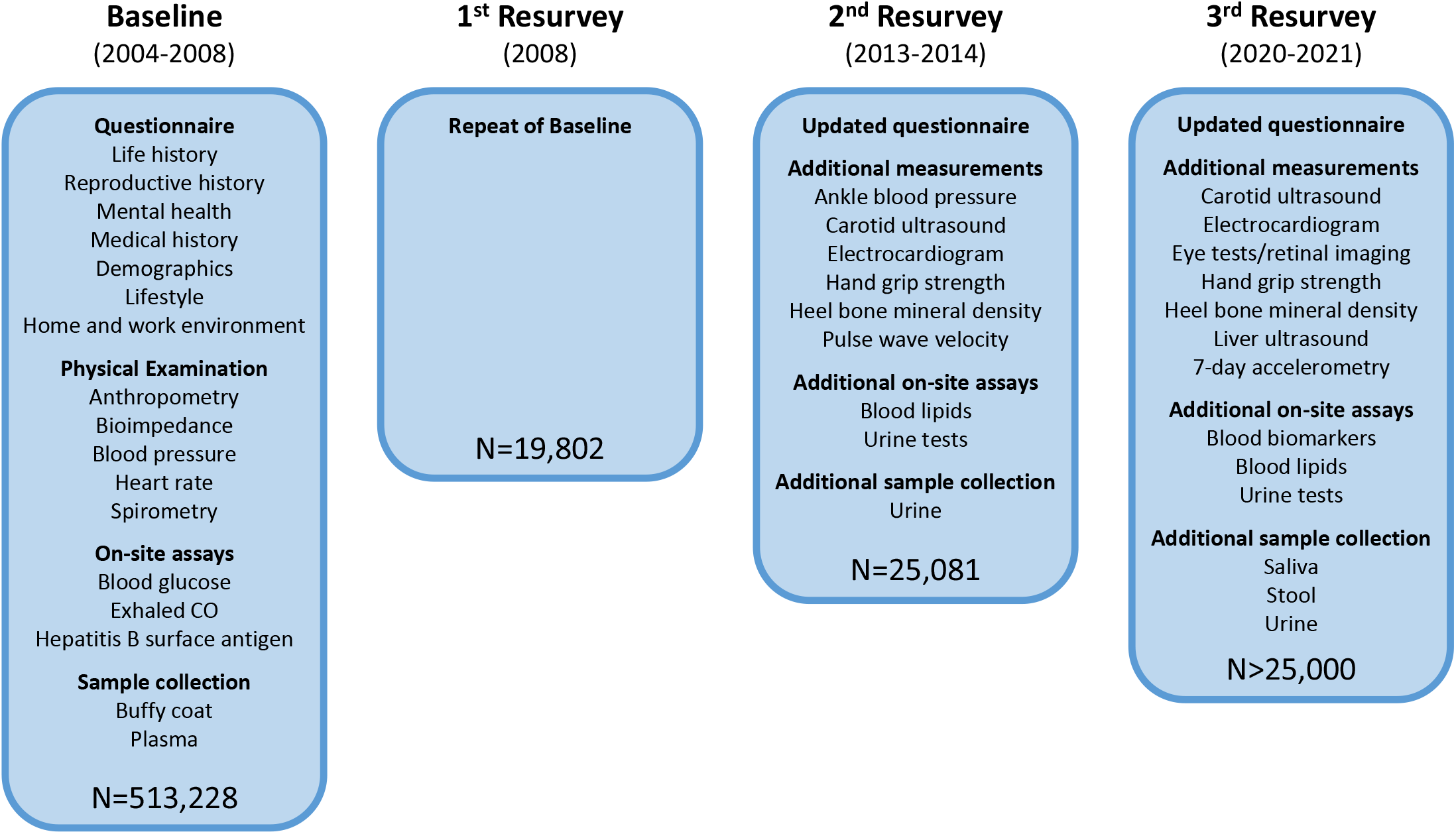
China Kadoorie Biobank survey details. Baseline questionnaire content, physical measurements, on-site assays, and biosample collection were repeated at three subsequent resurveys of approximately 5% of surviving participants. Second and third resurveys used an updated questionnaire, included additional physical measurements and on-site assays, and collected additional biosamples, as shown.

In addition to data collected at baseline and the resurveys, an increasing range of data is being generated from assays of stored biosamples. As part of a nested case-control study of stroke and ischaemic heart disease, plasma samples from up to 18,728 participants (all with genotyping) were assayed for 17 clinical biochemistry measurements, with ^1^H-NMR metabolomics for 4,657; further ^1^H-NMR metabolomics measurements were conducted for other nested case-control studies of pancreatic cancer and diabetes (2,500 samples to date). Initial assays using Olink proteomics (3,072 proteins) and Somalogic proteomics (up to 7,000 proteins) for a further nested case-subcohort study of myocardial infarction are in progress (3,977 participants, all with genotyping), with additional larger-scale measurements planned in the near future. Other assays in progress include multiplex serology of antibodies to antigens from 19 pathogens (in 4,500 samples to date, with measurement in 40,000 cancer cases and controls underway) and ^1^H-NMR metabolomics of urine samples from 25,251 second resurvey participants.

### Array design

CKB genotyping used custom-designed arrays on the Affymetrix (now ThermoFisher) Axiom^®^ platform, with content selection based on similar overall principles to those used for the UK Biobank array design^15,16^, but adapted to optimise performance for individuals of East Asian ancestry. This addressed three high-level criteria: (1) maximisation of genome-wide coverage of common and low-frequency variation in individuals across the whole of China; (2) detection of important variants and of rare loss-of-function and other protein-coding variants that are present in Chinese populations; and (3) consistent performance across array designs and batches of arrays manufactured over an extended time period.

Using the UK Biobank probe list as the starting point for the CKB array design, this was then modified and extended, informed by allele frequency and sequence data for over 12,000 East Asians that were available to us in 2013 (**Supplementary Information, Supplementary Figure S1**). In brief, we: (i) removed variants identified as absent or at low frequency in East Asians; (ii) added specific variants confirmed as being present in East Asians, including loss-of-function, missense, and eQTL variants; (iii) constructed an East Asian-specific genome-wide grid that maximised imputation of both common (>5%) and low frequency (1-5%) variants; and (iv) included multiple copies of a series of degenerate probes for detection and classification of circulating hepatitis B (HBV) viral DNA. The resulting array design comprised 781,937 probesets assaying 700,701 variants, of which 354,399 were also present on the UK Biobank array (**Supplementary Figure S2, Supplementary Table S1**). The design included duplicate probesets, one for each strand, for 81,236 variants which did not have a validated assay on the Axiom platform.

The initial array design was revised on the basis of genotyping data from the first 100 plates (8,995 samples after QC) (**Supplementary Information, Supplementary Figure S3**). Failed, poor quality, or otherwise-uninformative monomorphic probesets were removed, along with the poorer-performing probes of each pair of duplicate probesets. Further specific content of interest (including further HBV probes and tags for variants that failed QC) were added to the design. Finally, variants were added to improve or restore genome-wide imputation coverage. **Figure 2** summarises the content of the final updated array, comprising 804,496 probesets assaying 803,030 variants, of which 340,562 are present on the UK Biobank array (**Supplementary Table S2**).

**Figure 2.**
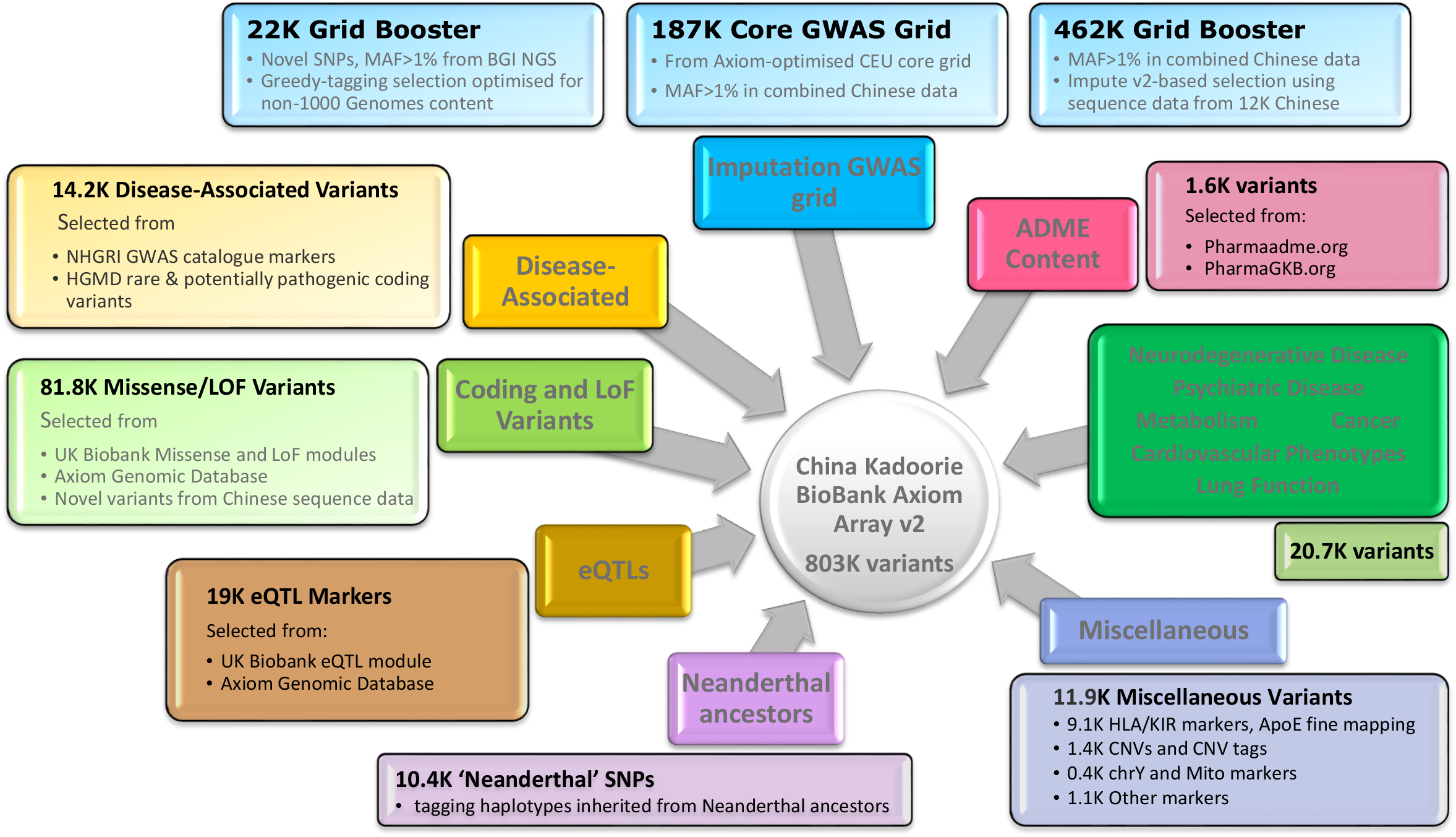
Design of the CKB Axiom^®^ genotyping array. The figure illustrates the different categories of content on the revised CKB array. Numbers indicate the approximate counts of variants in each category. Some variants fall into more than one category.

### Genotyping and quality control

Genotyping and quality control of a total of 105,408 CKB DNA samples are summarised in **Table 1**. The initial CKB array design was used to genotype 33,408 samples which had been selected for nested case-control studies of cardiovascular disease and COPD. Based on disease follow-up to 1 January 2014 (see below), this initial genotyping included all incident cases of intracranial haemorrhage (ICH: 5,020), subarachnoid haemorrhage (SAH: 455), and fatal ischaemic heart disease (fatal-IHD: 753); randomly selected incident cases of ischaemic stroke (IS: 5662), myocardial infarction (MI: 1008), and COPD (5376); participants with no cardiovascular events (n =10,038) at time of selection, matched to ICH cases for sex, age, and region; and 4,766 randomly-selected individuals who had attended the second resurvey.

**Table 1.** Genotyping sample selection and quality control.

|  | Array v1 | Array v2 | Total |
| --- | --- | --- | --- |
| **Total samples genotyped<sup>a</sup>** | **33,408** | **72,000** | **105,408** |
| Sample ascertainment |  |  |  |
| ICH | 5,020 | 602 | 5,622 |
| SAH | 455 | 46 | 501 |
| IS | 5,662 | - | 5,662 |
| MI | 1,008 | 1,028 | 2,036 |
| Fatal IHD | 753 | 163 | 936 |
| COPD hospitalisation | 5,358 | - | 5,358 |
| ICH-matched controls | 10,038 | - | 10,038 |
| Random selection | 4,766<sup>b</sup> | 69,378<sup>c</sup> | 74,144 |
| Intentional duplicates | 347 | 766 | 1,113 |
| Unintentional duplicates | 1 | 36 | 37 |
| **Total unique samples** | **33,060** | **71,198** | **104,277** |
| QC exclusions |  |  |  |
| Failed initial QC | 524 | 2,184 | 2,708 |
| Call rate <95% | 2 | 0 | 2 |
| Excess heterozygosity<sup>d</sup> | 89 | 253 | 342 |
| Excess homozygosity<sup>e</sup> | 3 | 0 | 3 |
| Sex mismatch | 47 | 121 | 168 |
| Other linkage error | 2 | 33 | 35 |
| XY aneuploidy<sup>f</sup> | 91 | 173 | 264 |
| Ancestry outlier | 3 | 1 | 4 |
| Consent missing/withdrawn | - | 31 | 31 |
| **Samples in current data release** | **32,300** | **68,406** | **100,706** |

<sup>a</sup> Excluding repeats of plate failures; <sup>b</sup> Random selection of samples from participants attending the second resurvey; <sup>c</sup> selected as complete boxes of DNA samples, prioritising boxes with large numbers of samples from participants eligible for the second resurvey; <sup>d</sup> >3 SDs greater than mean heterozygosity for participants recruited in the same region; <sup>e</sup> >3 SDs less than mean heterozygosity for participants recruited in the same region, and with total runs of homozygosity <2 SDs greater than the mean; <sup>f</sup> Identified as XXY, XY with non-negligible chrX heterozygosity, XXX, X0, or mosaic X0. Some samples failed QC on the basis of >1 criterion.

The updated array design was then used to genotype a further 72,000 samples, including all available additional cases of ICH (602), SAH (46), MI (1,028), and fatal-IHD (163) that had not previously been genotyped. The remaining genotyped samples came from boxes of DNA samples that were either randomly-selected or selected as containing samples derived from the clinics used for the second resurvey, in either case being largely representative of the overall CKB cohort. Duplicate samples were present on each pair of consecutive plates to support sample and plate tracking and for other quality-control purposes.

Genotyping and quality control followed the Affymetrix best practice workflow^17^ with additional checks for probesets that displayed substantial between-plate or between-batch differences in allele frequency or call rate. After QC and removal of duplicate probesets, 76.1% and 85.5% of probesets remained for the initial and revised arrays, respectively (**Supplementary Table S3**). 3.4% of samples failed QC (summarised in **Table 1**): these were mostly initial QC failures, likely reflecting low quality or low concentration DNA samples; only 0.16% of samples were excluded due to a sex mismatch, reflecting CKB’s stringent sample tracking procedures^18^. All pairs of duplicate samples showed high concordance of non-missing genotypes, overall concordance being 99.88% and 99.87% for the first and second array designs respectively (with 0.67% and 0.64% calls missing in one or both of a pair). Where probesets on the revised CKB array were present on the UK Biobank array, 192 samples genotyped on both arrays yielded concordance of 99.80% (0.46% missing).

The allele frequency distribution for the two datasets reflected the design characteristics of the arrays (**Figure 3A**). There were many more monomorphic or very low frequency (MAF<0.0001) variants on the original array design (5.7%) than on the revised array (1.6%) on which many such variants had been removed. Conversely, revision of the array design included selection of additional variants to improve imputation of variants with MAF of 0.01-0.05, and correspondingly more variants passed QC in this MAF range. The allele frequencies of variants that passed QC showed strong agreement with the East Asian populations in the 1000 genomes reference^19^ (**Figure 3B**), even at lower MAF where estimates in the 1000 genomes reference were impacted by low sample size, and this agreement was consistent across all recruitment regions, although with somewhat greater variation at lower MAF (**Supplementary Figure S4**).

**Figure 3.**
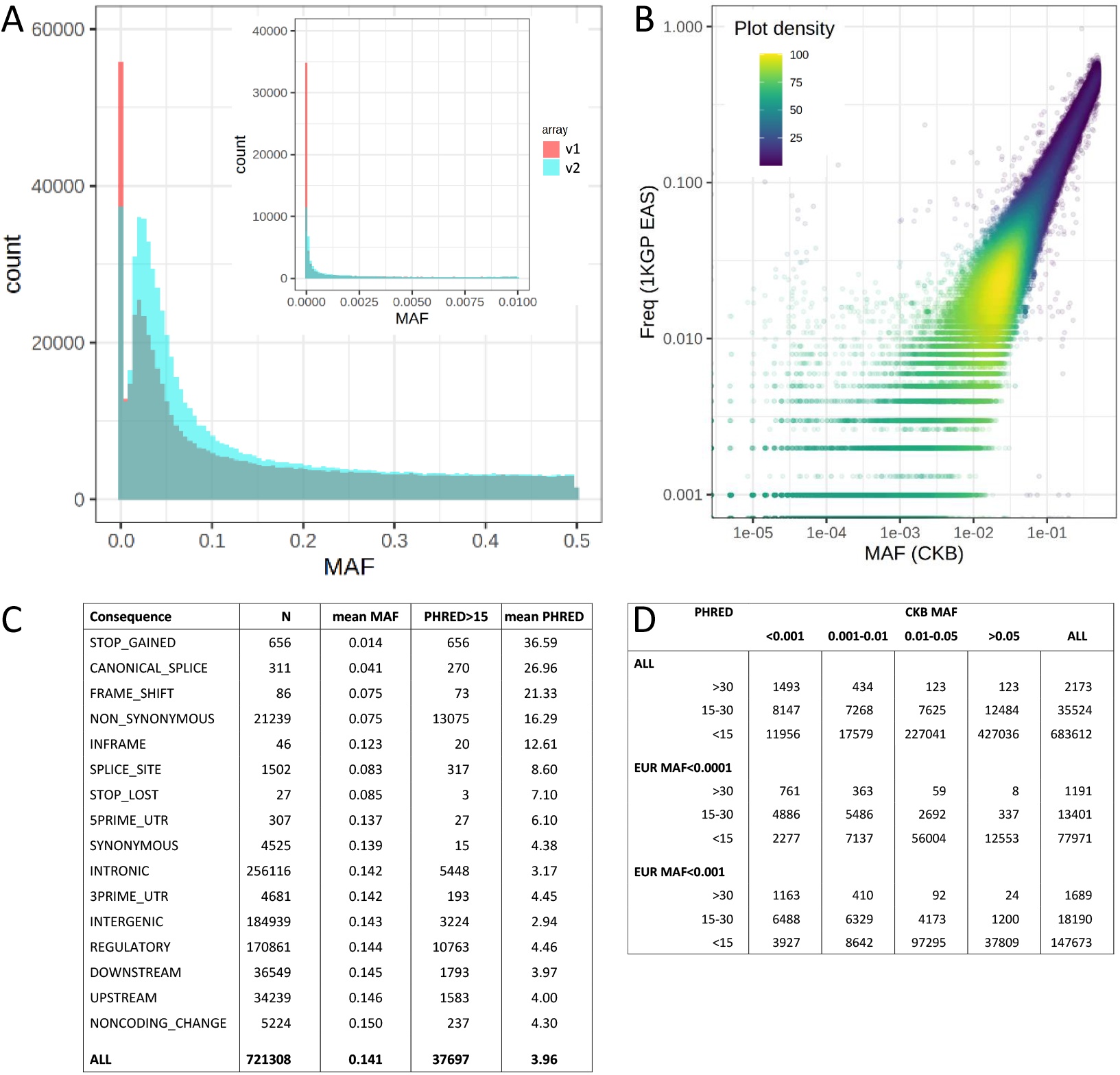
Allele frequency and functional annotation of genotyped variants. (A) Allele frequency distribution in unrelated CKB participants of variants passing QC on the two versions of the CKB genotyping array. (B) Comparison of CKB allele frequency of QCed variants on array v2 with the corresponding allele in the East Asian subset of the 1000 genomes Phase 3 reference. (C) Frequency and characteristics of different classes of QCed variants on array v2, according to Combined Annotation Dependent Depletion (CADD v1.6)^20,21^. (D) Allele frequency distribution in CKB and European populations of QCed variants on array v2, for variants with different levels of predicted functional impact according to CADD.

These allele frequency data provide some insight into the potential utility of variants included on the CKB array for purposes of investigating the impact of protein loss of function. Variants on the revised array that passed QC were categorised according to their potential functional significance as predicted by Combined Annotation Dependent Depletion (CADD v1.6)^20,21^; this identified 7 classes of variant annotation representing 23,867 variants which had both substantially lower mean MAF and higher mean PHRED than the other classes, indicating strong enrichment for deleterious variants (**Figure 3C**). Overall, 37,697 variants had a PHRED value >15, corresponding to the top 3% most damaging variants genome-wide, with a high likelihood of pathogenicity^22^. Of these variants, more than half (20,355, 54%) had a MAF >0.01 in CKB (28,057, 74% for CKB MAF>0.001), of which 5,489 (27%) are virtually absent from European populations (**Figure 3D**). These will provide opportunities not available in European cohorts for genetic investigations of the importance of the affected genes for disease and disease risk, such as those already conducted for *PLA2G7* and *CETP*^*5*,6^.

Initially, imputation was performed separately for each array dataset, but the revised array provided only a modest improvement in imputation quality, despite the substantially larger number of informative variants passing QC (**Supplementary Table S3**). Therefore, to minimise batch and array effects, we derived a single imputation dataset, using only those variants passing QC in all batches on both array versions (although variants excluded for imputation remain available for analysis in the final dataset). We achieved high confidence imputation for the large majority of common and low-frequency variants present in the EAS populations of the 1000 genomes Phase 3 reference (**Supplementary Tables S3-S4, Supplementary Figure S5**): Mean info score was 0.950 for variants with MAF>0.05, 0.849 for MAF 0.01-0.05, and 0.695 for MAF 0.005-0.01. Imputation was typically poorer for rare variants with MAF <0.005, which are excluded from many analyses.

### Relatedness

The community-based recruitment of CKB participants resulted in family groups attending together, so that many individuals had close relatives also recruited into the study (**Supplementary Tables S5-S6**). Among genotyped participants, 31.9% had an also-genotyped second-degree or closer relative (23.6% having at least one first-degree relative), with more relatedness in rural than in urban regions (39.0% vs 22.8% with first/second degree relatives), with the exception of participants in Suzhou (54.7%); Suzhou recruitment took place in a previously-rural district that has only recently become urbanised. Suzhou also had a particularly high proportion of individuals with multiple close relatives (1,561 [20%] with ≥3 genotyped relatives). Further analysis of relatedness in CKB also identified 32 pairs of twins, 13,875 individuals with at least 1 sibling (comprising 6,325 family groupings of up to 7 siblings) and 6,571 parent-child relationships, including 1,189 trios.

Several regions displayed patterns of relatedness suggestive of historical consanguinity (**Supplementary Figure S6**). Histograms of pairwise identity-by-descent (IBD) displayed not only the expected peaks corresponding to integer numbers of meioses separating relatives (at IBD of 0.5, 0.25, 0.125, etc.), but also other peaks centred on values corresponding to relationships which arise as a result of consanguineous unions between individuals with recent common ancestors, for instance triple second cousins (expected IBD of 0.09375). Consistent with this, 1050 participants had heterozygosity >3 SDs below the mean values across all genotyped samples, all but 3 of whom had correspondingly extensive runs of homozygosity (**Supplementary Figure S7**).

### CKB population structure

Principal component analysis (PCA) of 76,719 unrelated CKB participants identified 11 PCs informative for CKB population structure. Consistent with findings from many previous studies, individuals formed discrete clusters correlated strongly with longitude and latitude, mostly reflecting the geographic origins of the individuals recruited in each region (**Figure 4, Supplementary Figure S8**). For three regions, however, the clusters of participants were clearly offset on a plot of the first 2 PCs compared to their recruitment location. In each case the apparent discrepancies can be explained by known, relatively recent, major, historical population movements: large scale migration in the 16^th^ and 17^th^ centuries AD from Guangdong to Hainan island; repopulation of the Chengdu Basin in Sichuan in the late 17^th^ and early 18^th^ centuries, a large proportion of migrants coming from Huguang (now Hubei/Hunan); and settlement of largely-unpopulated Manchuria in the late 19th-early 20th century, with the majority of settlers originating from Shandong province.

**Figure 4.**
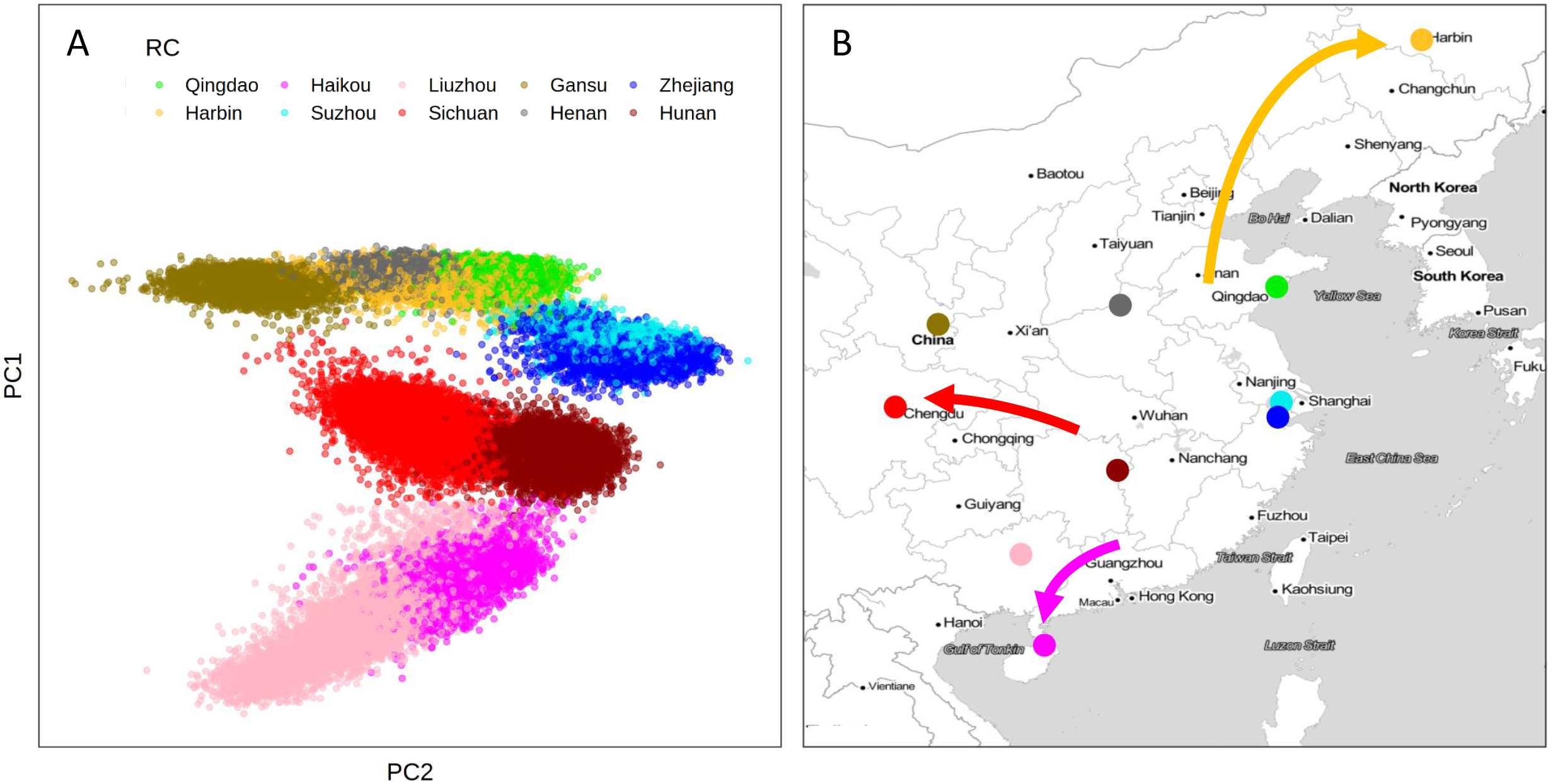
Correlation of CKB population structure with geographic origins. (A) Plot of the two leading principal components from PCA of CKB genotypes, with each participant colour-coded according to the regional centre (RC) where they were recruited. (B) Map of China and adjacent countries showing the locations of the ten CKB regional centres. Arrows denote major population movements in recent history that can account for mismatches in the correlation between PCA and geography.

Although the leading PCs tightly clustered most individuals for each recruitment region, a small proportion (5.7% overall) lay outside the main cluster (>3 SDs from the region mean for PCs 1-11) and appeared to have non-local ancestry. Of these, for those with data available from the second resurvey, a high proportion (47.8%) reported that they or at least one parent were born in a different province of China; by comparison, only 12.5% of the remainder reported origins from a province other than that in which they were recruited. The large majority of participants with non-local ancestry was recruited in Liuzhou, among whom a substantial fraction of individuals (25.4%) lay outside the main PCA cluster, mostly reflecting outlier values for PC1 (corresponding to the major north-south axis); 87.2% of these individuals reported that either they or a parent was born outside Guangxi province. Local population structure within each region was not readily apparent from the above pan-China PCA, but was clearly observed for individuals within each region-specific cluster (excluding those identified as having non-local ancestry) (**Figure 5, Supplementary Figures S9-S10**). Between 2 and 9 PCs were informative for the latitude and longitude of the study clinic at which an individual was recruited. Rural regions (plus previously-rural Suzhou) typically displayed substantial structure, reflecting established communities with little population movement; by contrast, there was only limited population structure for most urban regions. Once again, an exception was Liuzhou, for which an appreciable proportion of second resurvey participants reported having non-Han ancestry: Although uninformative for geographical location within Liuzhou, the first 4 PCs from whole-cohort PCA or the first 2 local PCs were informative for Han status (**Supplementary Figure S11**).

**Figure 5.**
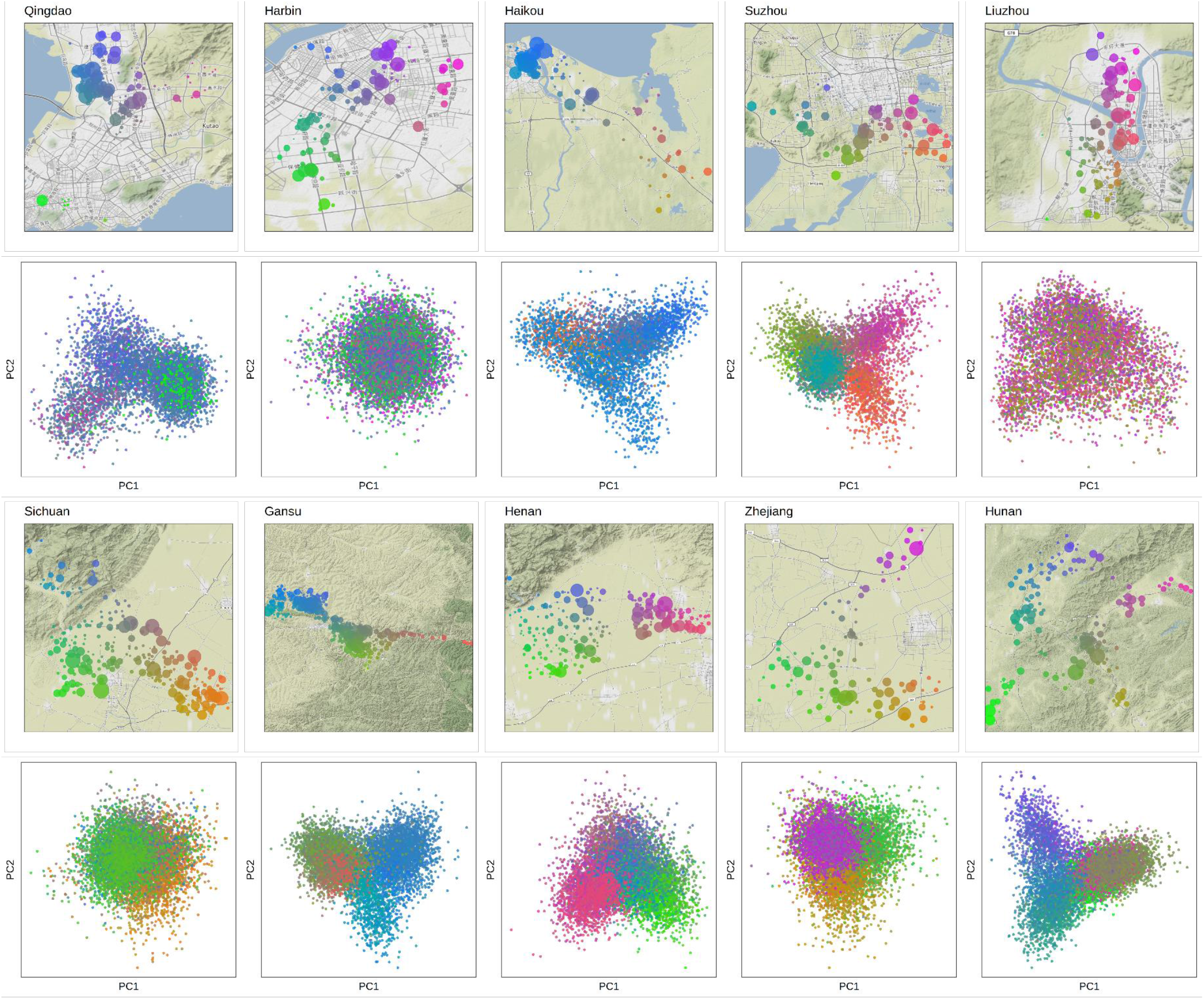
Local population structure in CKB regions. Local maps are shown for each recruitment region, showing the geolocation of the individual recruitment clinics, colour coded according to latitude and longitude; the size of the symbol is proportional to the number of genotyped individuals from that clinic. Corresponding PCA plots show the first two principal components from PCA of individuals from that region, colour coded according to their recruitment clinic. Top 2 rows – urban regions; bottom 2 rows – rural regions.

The geographical and/or historical relationships between the different CKB populations are also reflected in F_st_ analyses of the genetic distance between regions (**Supplementary Figure S12**): The four northern regions cluster together, and also with the Northern Han 1000 genomes population (CHB); the four regions situated on or near the Yangtze river cluster together, in two pairs, and also with the Southern Han 1000 genomes population (CHS); and the southern two regions cluster together, with no appreciable distinction in Liuzhou between Han and non-Han identity. The positions of the 1000 Genomes Project East Asian populations in this analysis indicate that the 10 regions in the CKB population are components of a continuum running from north to south with no clear separation from neighbouring countries (KHV, from Vietnam) or ethnic populations (Dai Chinese, from near to the borders with Laos and Myanmar).

### Disease outcomes

In common with the other biobanks contributing to the Global Biobank Meta-analysis initiative^14^, one of the chief strengths of CKB is the ability to follow up study participants for a wide range of fatal and non-fatal disease outcomes^2^. In CKB, disease follow-up is obtained by electronic linkage of participants’ unique national identity numbers to registries for death and for 4 major diseases (stroke, ischaemic heart disease, cancers, diabetes) and to the national health insurance system, which records all in-patient hospital events. These procedures are complemented by active follow-up through annual checks of local residential records and, if necessary, in-person visits by local staff to check key data and to identify hospitalised episodes in a small proportion of CKB (currently approximately 2%) who have not joined the health insurance scheme^23^. Data from these multiple sources, including parsing of free-text Chinese language disease descriptions and matching to a clinician-curated disease description library, are integrated and standardised into ICD-10 coded incident disease events. By 1 January 2019, >1.2M incident events and 49,428 deaths (including cause(s) of death) had been recorded for the whole of CKB, covering >5,000 separate disease types (defined according to three-character ICD-10 code), with only 5,302 (1.0%) of participants being lost to follow-up.

**Table 2** shows the number of individuals with selected incident events as recorded through disease follow-up, in all CKB participants and in the genotyped subset. Additional prevalent cases are available through medical questionnaire data, and on-site measurements at baseline (e.g. type 2 diabetes and COPD, through blood glucose assays and spirometry). Reflecting the strategy for selecting the genotyped samples, there was enrichment for cardiovascular diseases, including ischaemic stroke (ICD-10 code: I63), intracerebral haemorrhage (I61), and myocardial infarction (I21), for COPD (J41-J44, J47), and for all-cause mortality. By contrast, other diseases unrelated to the ascertained case types were present in proportion with the number of genotyped samples.

The wide range of disease outcomes recorded during follow-up is illustrated by the 225 different 3-character ICD-10 codes that have at least 100 incident events recorded in genotyped participants (**Supplementary Table S7**). Although limited, this number of cases is sufficient to permit analysis using software packages such as SAIGE^24^, and subsequent contributions to multi-cohort meta-analyses. Work is ongoing to convert the ICD-10 data into Phecodes^25^, to aid harmonisation of disease outcomes between CKB and other biobanks and to facilitate phenome-wide association analyses.

**Table 2.** Death and disease events in CKB. Numbers of CKB participants amongst the whole cohort and the genotyped subset who underwent selected death and disease events during follow-up prior to 1 January 2018.

| **Disease** | **Definition, ICD-10 codes** | **All participants** | **Genotyped** |
| --- | --- | --- | --- |
| Tuberculosis | A15-A19, J65, K23.0, K67.3, M01.1, M49.0, M90.0, N33.0, N74.0, N74.1 | 3,053 | 768 |
| Lung cancer | C33-C34 | 6,574 | 1,552 |
| Liver cancer | C22 | 3,256 | 665 |
| Stomach cancer | C16 | 3,771 | 756 |
| Diabetes | E10-E14 | 32,748 | 7,310 |
| MI | I21 | 7,984 | 3,386 |
| ICH | I61 | 11,638 | 6,663 |
| IS | I63 | 50,675 | 14,302 |
| Heart failure | I50 | 5,160 | 1,467 |
| Pneumonia | J12-J18 | 30,339 | 7,160 |
| Asthma | J45-J46 | 2,846 | 880 |
| COPD | J41-J44 | 20,391 | 7,971 |
| Liver cirrhosis | K70, K74 | 2,785 | 552 |
| Chronic kidney disease | N02-N03, N07, N11, N18 | 2,879 | 678 |
| Self-harm | T39.0, T39.1, T39.3, T40, T42, T43, T51, T52, T60, X60-X84, Y87.0, Z91.5 | 654 | 114 |
| **All hospitalised events** | A00-Z99 | **286,167** | **60,558** |
| **All deaths** | A00-Z99, underlying cause | **49,428** | **16,101** |

### Analytical approach

The historical consanguinity and extensive relatedness in CKB have been exploited in analysis of the impact of inbreeding on reproductive success^26^, and for within-sibship GWAS to derive estimates of direct genetic effects unaffected by genetic nurture^27^. However, for the majority of studies, the population structure of the CKB cohort together with the strategy for selection of samples for genotyping require thoughtful analytical approaches. The substantial relatedness within CKB, as in many other population-based cohorts, means that exclusion of individuals to avoid inclusion of pairs of close relatives (typically kinship >0.05, corresponding to third-degree relatives, e.g. first cousins) would result in a substantial reduction in sample size, with some recruitment regions being disproportionately affected (**Supplementary Tables S5, S6, S8**). Therefore, we typically use well-established software packages such as BOLT-LMM^28^ and SAIGE^24^ that implement linear mixed models to account for both relatedness and population stratification, thereby permitting inclusion of related individuals.

It is unclear, however, that current software packages always account for all aspects of population structure in CKB, with its recruitment in 10 discrete regions each with their own distinct genetic characteristics, varying environments, cultures, demographics, and incidence rates of major disease outcomes. For some diseases, this may be not only because they vary in prevalence but also because of varying access to healthcare (e.g. in rural compared to urban regions), so that patterns of severity in reported cases may also vary between regions. Therefore, while it is frequently appropriate to conduct analyses across the full genotyped dataset with adjustment for recruitment centre (by inclusion of 9 binary covariates), wherever possible we supplement this with region-stratified analyses (excluding the 5.7% of individuals with non-local ancestry) and meta-analysis, to ensure that association signals are not due to unresolved population stratification or subject to other biases (e.g. arising from heterogeneity between regions).

A second consideration is that the selection for genotyping of nested case-control samples has resulted in substantial over-representation of participants with hospitalisation for cardiovascular disease or COPD; although the additional cases were selected on the basis of incident events (i.e. after recruitment), the baseline characteristics of these individuals nevertheless differ from the overall population (e.g. cardiovascular disease events are positively associated with blood pressure, adiposity, blood lipids, smoking, and alcohol consumption). Their inclusion potentially introduces biases or confounding into analyses using the complete genotyping dataset, and we have therefore developed approaches that seek to minimise or eliminate these biases. For quantitative traits available for all participants, such as blood pressure or reproductive traits, we perform all adjustments for covariates and data transformations in the full CKB cohort, prior to genetic analyses, so that these adjustments are not distorted by the non-random nature of those genotyped; this is typically performed as a single regression including region as covariates, but we also check the impact of instead performing such adjustments in each region separately. Where traits are only available in non-random subsets of individuals (e.g. clinical biochemistry measures), we either exclude ascertained cases entirely, include case ascertainment as a covariate, or conduct analyses stratified by ascertainment – note that this is not required for measurements taken at the second resurvey, which was representative of surviving CKB participants.

For the analysis of dichotomous disease outcomes, to overcome these potential biases we have constructed a subset of 77,176 individuals representative of the full CKB cohort in which over-representation of ascertained disease cases was eliminated (**Supplementary Table S8, Supplementary Methods**). Analyses of disease outcomes and other binary phenotypes, including contributions to the first round of GBMI studies^14,29-31^ have typically used this population-representative subset supplemented with additional cases from the remainder of the dataset. For these analyses we use SAIGE software^24^, which is designed to account for imbalances in numbers of cases and controls. Where numbers of cases permit, this approach is combined with region-stratification and meta-analysis. It should be noted that use of the population subset does not result in a noticeable loss of power, despite the exclusion of ∼25% of samples, since there is invariably a large excess of controls even for more common diseases. It also provides a reduced dataset, unaffected by disease ascertainment biases, which can be used for sensitivity analyses of studies of other traits that use the full genotyped dataset.

### Research Contributions

In combination with genotyping and imputation, and the wide range of phenotypes and disease outcomes available for CKB participants, the above analytical approaches have been applied in diverse studies. Initial studies, using directly-genotyped variants, were MR-based investigations which emphasised the value of ancestry diversity for genetic analyses. In early examples of “drug target MR”, we found no association of East Asian-specific variants in *PLA2G7* and *CETP* with major cardiovascular events or other major diseases, in each case complementing the results of clinical trials which found no major benefit of drug treatments targeting their respective protein products^5,6,32^. We also exploited the high frequency in East Asians of variants influencing alcohol metabolism, and thereby drinking behaviour, to investigate the causal relationship between alcohol consumption and deleterious effects on health: we showed a clear link between alcohol and risk of stroke and, for the first time, we robustly refuted previous reports from observational studies of apparent protective effects of moderate drinking^33^. Other early genetic studies in CKB investigated the causal relevance of other disease risk factors, providing evidence that vitamin D deficiency increases risk of diabetes and cardiovascular disease^34,35^, that diabetes is itself causally associated with increased risk of cardiovascular disease^36^, and that, while lowering of LDL-cholesterol decreases risk of ischaemic stroke, it also increases risk of haemorrhagic stroke^37,38^.

Since becoming available, the genome-wide imputed data have enabled a wider range of studies, including MR investigation of further drug targets^7,8^ and of diverse disease risk factors such as bone mineral density^39^, gallstone disease^40^, and resting heart rate^41,42^. We have contributed to replication of novel association signals from large GWAS of blood pressure^43^, menopause^12^, and early-onset stroke^44^, and have evaluated the performance of polygenic scores in predicting disease risk for lung function^9,45^, lung cancer^46^, fracture^47^, and breast cancer^48^. A comprehensive set of GWAS for major diseases and disease risk factors are in progress: we recently published the first large GWAS of lung function in an East Asian population^49^, identifying 48 independent associations of which 18 were novel, once again emphasising the value of expanding ancestry diversity in genetic studies.

In addition to these analyses conducted primarily within CKB, we have contributed to many genome wide association studies in collaboration with major consortia, including trans-ancestry studies of intracranial aneurysm^10^, recurrent miscarriage^11^, blood lipids^50^, fingerprint patterns^51^, diabetes^52^, and height^13^. In particular, CKB has made important contributions to the growing number of studies focussed specifically on populations of East Asian ancestry, including the largest single contribution to a GWAS of depression in East Asians^53^; and a major contribution to a GWAS of type 2 diabetes, the largest East Asian GWAS to date^54^. Summary statistics from CKB GWAS have also contributed to the development of methods for: genetic association analyses using very-low-coverage whole genome sequencing from non-invasive prenatal testing^55^; trans-ancestry colocalisation to assess whether two populations share causal variants^56^; and improved genetic discovery in multi-ancestry meta-analyses^57^.

### Future Prospects

With its breadth of phenotypes and disease outcomes, prospective study design, and growing range of diverse ‘omics assays, CKB will continue to make significant contributions to genetic discovery and elucidation of disease aetiology and causality. Ongoing work will further enhance the available genetic resources, including DNA methylation arrays for 982 samples^58^; imputation using the Trans-Omics for Precision Medicine (TopMED)^59^ and Westlake Biobank for Chinese (WBBC)^60^ reference panels; and whole genome sequencing of 10,000 participants with incident ischaemic stroke (with preliminary analyses underway). Whole genome sequencing of the entire 512,000 cohort is planned in the near future through private-public partnership. Together with other notable biobanks across the world, CKB is addressing the recognised need for ancestrally diverse biobanks, and will continue to make strong contributions to the East Asian and trans-ancestry genetic analyses which are beginning to correct the strong Euro-centric bias of the genetic literature^4^.

## Methods

### Study permissions

All participants provided written informed consent at each survey visit, allowing access to their medical records and long-term storage of biosamples for future unspecified medical research purposes, without any feedback of results to the individuals concerned. Ethical approval was obtained from the Oxford Tropical Research Ethics Committee, the Ethical Review Committees of the Chinese Centre for Disease Control and Prevention, Chinese Academy of Medical Sciences, and the Institutional Review Board (IRB) at Peking University. The Chinese Ministry of Health approved the study at the start in 2004 (including export of plasma samples to Oxford), and also approved electronic linkage to health insurance records in 2011. Raw genotyping data were exported from China to the Oxford CKB International Coordinating Centre under Data Export Approvals 2014-13 and 2015-39 from the Office of Chinese Human Genetic Resource Administration.

### CKB study data

Full details of the CKB study design and methods have been previously reported^2^. Briefly, 512,726 adults aged 30-79 years were enrolled during 2004-2008 from ten urban and rural areas across China. At local study assessment clinics, trained health workers administered a laptop-based questionnaire; undertook physical measurements; and collected a blood sample for long-term storage and onsite blood tests. Three subsequent resurveys of ∼5% randomly selected surviving participants were conducted using similar procedures in 2008, 2013-2014, and 2020-2021. With the exception of genomics data, all CKB survey data were collected and stored using bespoke IT systems and databases tailored to CKB requirements^61,62^. Disease follow-up data from death and disease registries and from health insurance records were processed and matched to study identifiers by local staff in each recruitment region, centrally processed and converted into ICD-10-coded events, and integrated into the main study database. The database is regularly processed into research-ready snapshots from which datasets are served to researchers. All results are based on CKB data release version 17.02, incorporating disease follow-up up to 1 January 2019.

### DNA extraction and genotyping

DNA extraction and genotyping was performed at BGI, Shenzhen, China, and data processing, genotype calling, and quality control was conducted at CTSU, University of Oxford, UK (see **Supplementary Methods** for full details). DNA was extracted from 800µL stored buffy coat using KingFisher™ Blood DNA Kit and KingFisher™ Flex 24 Magnetic Particle Processors (Thermo Scientific), yielding 400µL DNA at 220 ng/L mean concentration. The first 95,680 DNA samples (from randomly-selected participants), extracted during 2012-2013, were genotyped using the multiplex Golden Gate^®^ platform (Illumina), for panels of 384 variants which included 3 variants informative for sex within the chromosome XY pseudoautosomal regions. Duplicate samples had overall genotyping concordance of 99.98%, and 2.5% of samples failed quality control, with only 0.1% of samples with a sex mismatch and 0.1% with other potential sample linkage errors, confirming good DNA quality and robust linkage to originating study participants.

DNA samples selected for genome-wide genotyping were sub-aliquoted and diluted to a consistent concentration of 50ng/µL as measured by Qubit DNA quantification (ThermoFisher), and genotyping was performed with manual target preparation according to Affymetrix protocols with automated plate processing and imaging using CKB_1 and CKB_2 Axiom^®^ arrays and GeneTitan^®^ Instruments^63^. Genotyping quality control and calling was performed according to Affymetrix Best Practice workflow^17^ using the Axiom Analysis Suite (Affymetrix) with default settings. After exclusion of plates failing initial quality metrics and of individual samples with call rate <97% for 20,000 high performance variants, batches of 50 plates were processed and co-clustered to derive genotypes and further quality metrics. Batch level QC identified probesets with poor clustering, call rate <95%, or with between-plate or between-batch effects; probesets failing QC in any batch or displaying significant batch effects were excluded from the final dataset. After performing preliminary sample QC, probesets were excluded with overall call rate <98%, deviation from Hardy-Weinberg equilibrium, or minor allele frequency >0.2 different from that for the 3 Chinese populations from the 1000 Genome Project Phase 3 reference^19^. Finally, for variants were assayed by 2 different probesets both passing QC, the probeset with the lower overall call rate was excluded. Overall performance of the revised array was tested using 192 samples (152 Chinese, 40 European) from the European Vasculitis Genetics Consortium^64^ genotyped using both CKB_2 and UK Biobank Axiom^®^ arrays. Combined Annotation Dependent Depletion (CADD v1.6)^20,21^ was used to look up the predicted functional consequences of 721308 variants passing QC on the CKB_2 array; the corresponding allele frequency in Europeans was according to the dbGAP Allele Frequency Aggregator (ALFA)^65^ v2020-11-14, population SAMN10492695.

Sample QC was on the basis of criteria as summarised in **Table 1**. Based on genotyped variants passing QC as above, samples were excluded from the dataset if they had overall cate rate <95%, excess heterozygosity (Z-score >+3), or excess homozygosity (Z-score <–3) not accounted for by extensive runs of homozygosity (**Supplementary Figure S7**). Samples from individuals with appreciable non-Chinese ancestry was identified from outlying values (>10 SDs from the mean) after projecting onto the first 10 principal components from 2,504 individuals from 26 populations (5 ancestries) from the 1000 genomes project (**Supplementary Figure S13**). Initial mismatches of computed sex with that recorded in the CKB database were identified using chromosome X heterozygosity; all samples within blocks of sex-mismatched samples were excluded from analyses. Further checks for sex mismatch and for chromosome XY aneuploidies used plots of chrY/chrX probe intensity ratio against chromosome X heterozygosity, combined with visual checks of chrX heterozygosity for samples with outlying values for mean chrX probe intensity (**Supplementary Figure S14**).

### Imputation

Prior to imputation, additional QC excluded variants at multiallelic sites or with mismatched alleles compared with the reference. Imputation was conducted for each array version separately, in each case excluding variants that failed QC in any genotyping batch, and for a combined dataset limited to variants passing QC on both array versions. Phasing and imputation used biobank-scale SHAPEIT3 and IMPUTE4 software^16,66^ for autosomes, and earlier SHAPEIT2 and IMPUTE2 versions for chromosome X. Imputation used the 1000 Genomes Project Phase 3 reference^19^ filtered to exclude variants with MAF=0 in the 5 East Asian populations. Post-imputation checks excluded variants displaying significant batch/array effects.

### Genetic analyses

Unless otherwise specified, genetic analyses were conducted using PLINK v1.9 and PLINK v2.0^67^. Sets of unrelated samples for variant QC and F_st_ analyses were derived using the --rel-cutoff option, while PCA and exclusions for GWAS used --king-cutoff 0.05, in each case determined using LD-pruned sets of 122,675 autosomal variants derived using the --indep-pairwise option. Relatedness between pairs of individuals used PI-HAT, Z0 and Z1 from identify-by-descent using the --genome gz option; first and second degree relatives were defined using PI-HAT thresholds of >0.375 and >0.1875, respectively. Parent-child pairs were identified as those with Z0 <0.05 and Z1 >0.5.

PCA was conducted using FlashPCA v2.1^68^ after LD pruning and exclusion of regions of long-range linkage disequilibrium (LD); these were identified as sets of nearby variants contributing disproportionately to region-informative PCs (**Supplementary Figure S15**), using an approach similar to that previously used for UK Biobank^69^ except that the regions were identified using a hidden Markov chain model incorporating between-variant recombination rates. A total of 223 regions were identified (**Supplementary Table S9**) and variants within these regions were excluded from the set used for further PCA analysis so that 171,236 variants and 76,719 unrelated CKB participants were included in the PCA; PCs for the remaining individuals were derived from the corresponding variant weights. PCs were determined to be informative for CKB population structure on the basis of the Bayes Information Criterion (BIC) for models predicting individuals’ recruitment region (**Supplementary Figure S16**), confirmed by ANOVA tests for non-random association of PCs with region of recruitment; above-trend eigenvalues on a scree plot; and visual examination of plots of the top PCs with colour-coding of region of recruitment (**Supplementary Figure S8**). Similarly, PCs informative for local population structure were identified on the basis of BIC for models predicting latitude and longitude for individuals’ recruitment clinic (**Supplementary Figure S10**) or Han status (**Supplementary Figure S11**). Individuals with appreciable ancestry not local to their recruitment region were identified from a robust Mahalanobis distance of >3 SDs for one or more of the informative PCs for that region. Maps used in PCA plots were drawn with R package ‘ggmap’^70^ using map tiles by Stamen Design (maps.stamen.com) under CC BY 3.0, using data by OpenStreetMap under Open Data Commons Open Database License.

### CKB Data Access

Data from baseline, first and second resurveys, and disease follow-up are available under the CKB Open Access Data Policy to bona fide researchers. Sharing of genotyping data is currently constrained by the Administrative Regulations on Human Genetic Resources of the People’s Republic of China. Access to these and certain other data is available through collaboration with CKB researchers. Full details of the CKB Data Sharing Policy are available at www.ckbiobank.org.

## Author Contributions

Data collection and analysis – RGW, IYM, KL, DSV, PM, AH, DA, NA, WWK, MAA, PAL, XJ, CN, YG, CY; funding – RGW, IYM, RC, PD, MRH, RP, HS, XX, RJC, LL, ZC; study oversight: RGW, RC, RP, JL, RJC, LL, ZM; manuscript draft and revision – RGW, IYM, KL, RJC, ZC. All authors reviewed the submitted manuscript.

## Acknowledgements

The most important acknowledgement is to the participants in the study and the members of the survey teams in each of the 10 regional centres, and to the project development and management teams based at Beijing, Oxford and the 10 regional centres. China’s National Health Insurance provides electronic linkage to all hospital treatments. We thank Judith Mackay in Hong Kong; Yu Wang, Gonghuan Yang, Zhengfu Qiang, Lin Feng, Maigeng Zhou, Wenhua Zhao, Yan Zhang, and Zheng Bian in China CDC; Lingzhi Kong, Xiucheng Yu, and Kun Li in the Chinese Ministry of Health; and Garry Lancaster, Sarah Clark, Martin Radley, Mike Hill, Hongchao Pan, and Jill Boreham in the CTSU, Oxford, for assisting with the design, planning, organisation, and conduct of the study. We thank Jeanette Schmidt, Teresa Webster, Yontao Lu, and colleagues at Affymetrix, Santa Clara CA, USA, for genotyping array design; Rory Bowden for helpful discussions during design of HBV probes included on the genotyping array; and Hongcheng Zhou, Haoxiang Lin, Jieqin Liang, and their colleagues at BGI, Shenzhen, China for DNA extraction, genotyping array design, and genotyping. P.A.L. would like to acknowledge financial support from Versus Arthritis (20593) and the British Heart Foundation (PG/13/64/30435) and to thank Cambridge Genomic Services (Cambridge, UK) for genotyping support, and we acknowledge the European Vasculitis Genetics Consortium for access to data. The CKB baseline survey and the first re-survey were supported by the Kadoorie Charitable Foundation in Hong Kong. Long-term follow-up was supported by the Wellcome Trust (212946/Z/18/Z, 202922/Z/16/Z, 104085/Z/14/Z, 088158/Z/09/Z), the National Key Research and Development Program of China (2016YFC0900500, 2016YFC0900501, 2016YFC0900504, 2016YFC1303904), and the National Natural Science Foundation of China (91843302). DNA extraction and genotyping was funded by GlaxoSmithKline, and the UK Medical Research Council (MC-PC-13049, MC-PC-14135). The project is supported by core funding from the UK Medical Research Council (MC_UU_00017/1, MC_UU_12026/2, MC_U137686851), Cancer Research UK (C16077/A29186; C500/A16896), and the British Heart Foundation (CH/1996001/9454) to the Clinical Trial Service Unit and Epidemiological Studies Unit and to the MRC Population Health Research Unit at Oxford University. Computation used the Oxford Biomedical Research Computing (BMRC) facility, a joint development between the Wellcome Centre for Human Genetics and the Big Data Institute supported by Health Data Research UK and the NIHR Oxford Biomedical Research Centre; the views expressed are those of the authors and not necessarily those of the NHS, the NIHR, or the Department of Health. We are grateful to Ida Surakka and Cristen Willer for helpful comments during drafting of the manuscript.

## License

This research was funded in whole, or in part, by the Wellcome Trust [212946/Z/18/Z, 202922/Z/16/Z, 104085/Z/14/Z, 088158/Z/09/Z]. For the purpose of Open Access, the author has applied a CC-BY public copyright licence to any Author Accepted Manuscript version arising from this submission

## Supplementary Information

Supplementary Information (CKB Collaborative Group, CKB Array Design, CKB Array Revision, Supplementary Methods)

Supplementary Tables S1-S9

Supplementary Figures S1-S15

## Notes

### Competing Interest Statement

The authors have declared no competing interest.

### Author Declarations

Oxford Tropical Research Ethics Committee of University of Oxford gave ethical approval for this work. Ethical Review Committees of the Chinese Centre for Disease Control and Prevention, Chinese Academy of Medical Sciences gave ethical approval for this work. Institutional Review Board (IRB) at Peking University gave ethical approval for this work.

