## Supplementary Information for "Genotyping and population structure of the China Kadoorie Biobank"

**Supplementary Information** **Page**

| | |
|---|---|
| Members of the China Kadoorie Biobank Collaborative Group | 2 |
| China Kadoorie Biobank Array Design | 3 |
| China Kadoorie Biobank Array Revision | 8 |
| Supplementary Methods | 13 |
| References | 20 |

### Members of the China Kadoorie Biobank Collaborative Group

**International Steering Committee:** Junshi Chen, Zhengming Chen (PI), Robert Clarke, Rory Collins, Yu Guo, Liming Li (PI), Chen Wang, Jun Lv, Richard Peto, Robin Walters.

**International Co-ordinating Centre, Oxford:** Daniel Avery, Derrick Bennett, Ruth Boxall, Ka Hung Chan, Yumei Chang, Yiping Chen, Zhengming Chen, Johnathan Clarke; Robert Clarke, Huaidong Du, Zammy Fairhurst-Hunter, Hannah Fry, Simon Gilbert, Alex Hacker, Mike Hill, Michael Holmes, Pek Kei Im, Andri Iona, Maria Kakkoura, Christiana Kartsonaki, Rene Kerosi, Kuang Lin, Mohsen Mazidi, Iona Millwood, Qunhua Nie, Alfred Pozarickij, Paul Ryder, Saredo Said, Sam Sansome, Dan Schmidt, Paul Sherliker, Rajani Sohoni, Becky Stevens, Iain Turnbull, Robin Walters, Lin Wang, Neil Wright, Ling Yang, Xiaoming Yang, Pang Yao.

**National Co-ordinating Centre, Beijing:** Yu Guo, Xiao Han, Can Hou, Chun Li, Chao Liu, Jun Lv, Pei Pei, Canqing Yu.

**Regional Co-ordinating Centres:**

**Gansu:** Gansu Provincial CDC – Caixia Dong, Pengfei Ge, Xiaolan Ren. Maiji CDC – Zhongxiao Li, Enke Mao, Tao Wang, Hui Zhang, Xi Zhang. **Haikou:** Hainan Provincial CDC – Jinyan Chen, Ximin Hu, Xiaohuan Wang. Meilan CDC – Zhendong Guo, Huimei Li, Yilei Li, Min Weng, Shukuan Wu. **Harbin:** Heilongjiang Provincial CDC – Shichun Yan, Mingyuan Zou, Xue Zhou. Nangang CDC – Ziyan Guo, Quan Kang, Yanjie Li, Bo Yu, Qinai Xu. **Henan:** Henan Provincial CDC – Liang Chang, Lei Fan, Shixian Feng, Ding Zhang, Gang Zhou. Huixian CDC – Yulian Gao, Tianyou He, Pan He, Chen Hu, Huarong Sun, Xukui Zhang. **Hunan:** Hunan Provincial CDC – Biyun Chen, Zhongxi Fu, Yuelong Huang, Huilin Liu, Qiaohua Xu, Li Yin. Liuyang CDC – Huajun Long, Xin Xu, Hao Zhang, Libo Zhang. **Liuzhou:** Guangxi Provincial CDC – Naying Chen, Duo Liu, Zhenzhu Tang. Liuzhou CDC – Ningyu Chen, Qilian Jiang, Jian Lan, Mingqiang Li, Yun Liu, Fanwen Meng, Jinhuai Meng, Rong Pan, Yulu Qin, Ping Wang, Sisi Wang, Liuping Wei, Liyuan Zhou. **Qingdao:** Qingdao CDC – Liang Cheng, Ranran Du, Ruqin Gao, Feifei Li, Shanpeng Li, Yongmei Liu, Feng Ning, Zengchang Pang, Xiaohui Sun, Xiaocao Tian, Shaojie Wang, Yaoming Zhai, Hua Zhang, Licang CDC – Wei Hou, Silu Lv, Junzheng Wang. **Sichuan:** Sichuan Provincial CDC – Xiaofang Chen, Xianping Wu, Ningmei Zhang, Weiwei Zhou. Pengzhou CDC – Xiaofang Chen, Jianguo Li, Jiaqiu Liu, Guojin Luo, Qiang Sun, Xunfu Zhong. **Suzhou:** Jiangsu Provincial CDC – Jian Su, Ran Tao, Ming Wu, Jie Yang, Jinyi Zhou, Yonglin Zhou. Suzhou CDC – Yihe Hu, Yujie Hua, Jianrong Jin Fang Liu, Jingchao Liu, Yan Lu, Liangcai Ma, Aiyu Tang, Jun Zhang. **Zhejiang**: Zhejiang Provincial CDC – Weiwei Gong, Ruying Hu, Hao Wang, Meng Wang, Min Yu. Tongxiang CDC – Lingli Chen, Qijun Gu, Dongxia Pan, Chunmei Wang, Kaixu Xie, Xiaoyi Zhang.

### China Kadoorie Biobank Array Design

The overall scheme for design of the CKB array is shown in **Figure S1**. Array content was selected from 8 distinct (but overlapping) classes:

1. Variants specified for various purposes by the CKB study group and collaborators;
2. Known GWAS hits present in the GWAS Catalog plus additional loci provided by colleagues and collaborators prior to publication;
3. Putative 'functional' variants identified in BGI sequencing data;
4. Content from (non-GWAS) modules defined for the UK Biobank array design;
5. Content from (non-GWAS) modules from Affymetrix catalogue arrays;
6. The optimised CEU core GWAS module, as used in the UK Biobank array;
7. ASN (JPT/CHB/CHS) SNPs/indels identified by HapMap/1000 genomes;
8. SNPs/indels with MAF>0.01 in unpublished sequence data (BGI).
9. Viral sequences for detection of Hepatitis B virus infection/subtypes;

The data sources used to select content included:

a. 1000 genomes Phase I data from 197 CHB/CHS subjects
b. 1000 genomes pilot data from 91 CHB subjects (for SNPs absent from the Phase I data)
c. BGI high coverage WGS data for 156 of the CHS/CHB 1000genomes subjects
d. BGI WGS data from 1,746 exomes from subjects mainly from southern China
e. Allele frequency data from genotyping of 1,802 subjects using the Taiwan Biobank array
f. Low coverage sequencing data from ~9,000 Chinese from the CONVERGE consortium

Together, these were used (A) to define pre-specified content on the array; and (B) to build a Chinese-optimised GWAS grid.

For use in SNP selection (and for determining MAF-defined SNP target lists), allele frequencies from (f) were used where available. Otherwise, data from (a/b), (d), (e) were combined, except that the 1000 genomes data (a) were replaced by BGI WGS data (c) if the latter gave a non-zero number of minor alleles – these high-coverage data were expected to be more accurate than the 1000 genomes low-coverage data.

The principles underlying the array design included:

- Maximising overlap with the UK Biobank array
- Taking account of Chinese-specific content
- Ensuring detection of specific important variants
- Maximising space-efficiency of the selected variants

Thus, the UK Biobank design was taken as a starting point, with the default being to include UKB content unless there was a good reason not to do so.

#### *A. Pre-Specified Content*

Pre-specified content was determined as follows. The marker counts given are for those that survived probe design QC.

1. *Affymetrix CEU GWAS core*

The optimised CEU GWAS core (UK Biobank module ‘GWAS Grid’) contained 246,055 SNPs and indels. SNPs/indels were removed from this list if they had MAF<0.01 or were not present in the 1000 genomes Phase I data (used by Affymetrix for imputation aware SNP selection) and, therefore, were not able to contribute to building of the GWAS grid, leaving 191,056 SNPs. Some markers were subsequently added back due to being in other modules (see below) or during GWAS grid selection, so that in total 193,326 markers from this module were included on the array.

2. *Other UK Biobank non-GWAS module content*

Markers found in Chinese samples:

All SNPs on the UK Biobank array that had been specified for some reason other than GWAS were checked for their presence in Chinese populations. All variants identified in at least one individual in the datasets above were provisionally included. This included all variants in several UKB modules.

Markers not found in Chinese samples:

The remaining variants, for which no instance of the minor allele was identified in Chinese samples, were treated as follows:

‘HLA/KIR’, ‘KIR’, ‘chrMT’, ‘chrY’, ‘ApoE’, ‘CNV Coverage’, ‘CNV tag’, ‘Fingerprint’, ‘ADME’, ‘Blood’, ‘BP/HT’, ‘Neanderthal’, ‘Alzheimers’, ‘eQTL’, ’Lung Function’: A relatively small proportion of these modules were not found in the available Chinese data. It was decided to include all such variants on the array, irrespective of detection in Chinese.

‘Cancer’, ‘HGMD’, ‘cardiac’, ‘neuro’: 6 well-evidenced cancer-related SNPs common in CEU but not already included were included. The remaining markers not already selected and present only in these modules were excluded.

‘missense’ or ‘LOF’ modules. These were mainly low-frequency variants selected on the basis of detection in the UK population. Such variants not present in Chinese were excluded.

Altogether, a further 100,411 variants were added to the array design, giving a total of 293,737.

3. *Affymetrix modules*

Markers from the Affymetrix catalogue modules eQTL, Exome319 and LOF, that were not included on the UK Biobank design but were detected in one or more of our Chinese datasets, were added to the array. The additional markers totalled 33,546, giving 327,283 in total.

4. *Known GWAS hits*

The NHGRI catalogue was downloaded on 6 December 2013. 11,745 unique lead SNPs were identified. These were merged with the UK Biobank ‘GWAS compatibility’ module, which included some tag SNPs (i.e. some loci had both the original hit and a tag SNP) and unpublished GWAS loci, giving a total of 12,735 markers. All of these were included on the array (QC permitting), irrespective of their prior detection in Chinese populations. As a result, a further 5,626 SNPs were added to the array, giving 332,909 in total.

*5. CKB Collaborative Group-selected SNPs*

683 SNPs and indels were specified by the CKB group and/or collaborators – including all SNPs previously successfully genotyped on the Illumina Golden Gate platform in 100,000 subjects – many of which were already included on the array. As a result, a further 252 SNPs were added to the array, giving 333,161 in total.

*6. Functional and chrY/MT SNPs from BGI data*

BGI provided data for 10,662 coding SNPs novel to Chinese (many completely novel) with putative functional effects (missense and nonsense), identified in datasets (c) and (d), and also some novel chrY/MT variants. To avoid private variants or calls due to sequencing errors, those identified in 1-2 individuals were excluded. Novel chrY/MT variants from Taiwan Biobank data were also included. In total, 5,288 additional variants were included on the final array design, giving 338,449 in total.

*7. Detection of Hepatitis B virus infection and type*

The available aligned HBV sequence data in late 2013 were downloaded from https://hbvdb.ibcp.fr/HBVdb/HBVdbIndex and used to calculate the entropy – i.e. the amount of variation – for each site along the genome. A sliding window of 71bp was then used to calculate the average entropy along the sequences, to identify the most conserved regions suitable for probe design. Six regions of approximately 100bp were identified, one of which contained a series of sequence variants that were expected to be sufficient to distinguish between HBV Genotypes B and C (the most common in China). For each candidate probe region, sequence variation at each site was tallied. Sites with more than one variant with a frequency ≥0.01 were recorded as 'SNPs' which would be catered for during probe design. The frequencies of all other variants (i.e. with frequency <0.01) were summed and recorded as 'residual variation' at each site.

Probe design was carried out based on sites that could be treated as 2-allele SNPs for the purposes of array data analysis. 35-mers in each direction from such 'SNPs' were recorded, treating multiallele 'SNPs' within those probes as degenerate positions requiring the design of multiple probe sequences. For each candidate 35-mer, residual variation across its full length was summed, to give a parameter "risk of probe failure" – viruses with a lot of inter-individual residual variation in these regions would potentially remain undetected due to poor probe hybridisation. Different alternative probes for each candidate probe region were compared in terms of their degeneracy and risk of failure, and the location of each probe set was selected so as to minimise these parameters.

For the majority of probe regions, whose primary purpose was to provide 'yes/no' detection of HBV, non-overlapping probe sets were selected. However, for the region diagnostic for Genotypes B and C, multiple overlapping probesets were designed so as to ensure that all diagnostic variant sites were interrogated. 15 different probesets were designed, with degeneracy at sites with variants with frequency ≥0.01, giving a total of 123 unique sequences. These were confirmed as having no appreciable homology to the human genome. Each was present in 8 copies on the array, to improve the ability to reliably detect low copy-numbers of HBV DNA.

#### *B. GWAS Grid Selection*

Chinese-specific MAFs were estimated for each marker in latest available 1000 genomes content (Phase I) as noted above, using the available data sources. These were allocated to bins corresponding to 0.05≤MAF≤0.50 and 0.01≤MAF<0.05, representing the sets of target markers to be tagged by the GWAS grid. The candidate SNPs available for selection were the full set of CHB/CHS/JPT 1000 genomes content for which Axiom assays could be designed (i.e. taking account of potential nearby interfering variants and/or sequences with appreciable similarity to other regions of the genome).

##### *1. Selection of SNPs to tag 1000 genomes content (Affymetrix)*

Using the pre-selected markers as the starting point, the Affymetrix imputation-aware SNP selection procedure was applied, initially targeting the SNPs with MAF≥0.05. Where there was a choice of SNPs to add to the array design, SNPs were prioritised that met one or more of the following criteria:

- Axiom-validated
- Not A/T or G/C allele pairs (i.e. requiring less array 'real estate')
- Present on UK Biobank array

The imputation coverage (proportion of target markers imputable at $r^2 \geq 0.8$) for each chromosome was determined at the end of each phase, and sufficient SNPs were selected and added to the array design such as to achieve 93% coverage for each chromosome. This procedure resulted in the addition of 237,246 additional variants to the final array design.

Despite this high overall coverage, some chromosomal regions remained for which coverage was poor. These were visually identified by randomly sampling 80,000 target markers for each chromosome and plotting their imputation $r^2$ values against chromosomal coordinate. Coverage in these regions was improved by selecting a further 12,107 SNPs, giving a total of 587,802 markers.

##### *2. Selection of SNPs to tag novel Chinese content (BGI)*

The analysis of WGS data available to BGI that provided putative functional variants (see pre-specified content part 6) also identified numerous common and low-frequency SNPs and indels not reported in 1000 genomes CHB/CHS/JPT populations. Coverage of those not already tagged (by 1000 genomes markers that could already be imputed using the currently-selected markers) was achieved using a greedy-tagging procedure with SNP prioritisation on the basis of the same criteria as Affymetrix SNP selection, combined with a score calculated using the sequencing quality scores of tagged and tagging SNPs and the extent to which untagged variants would tag other nearby untagged variants (using pairwise LD calculated from BGI's data).

This was carried out first for tagging of SNPs with MAF≥0.05, with selection of ~60,000 SNPs, and then for tagging of SNPs with 0.01≤MAF<0.05, with selection of ~20,000 SNPs. The final number of additional SNPs selected was 80,370, giving a total of 668,172 markers.

##### *3. Selection of SNPs to increase coverage of low frequency 1000 genomes content (Affymetrix)*

To improve imputation coverage of low-frequency variants, the remaining capacity of the array was used for a further 32,529 SNPs, selected using the Affymetrix imputation aware procedure. This gave a total of 700,701 markers.

#### 4. *Final QC and array design confirmation*

During final array design, a number of markers were identified for which it was not possible to design array probes or for which pairs of probes were sufficiently similar in sequence that it was necessary to exclude one of them. Where this occurred, alternative SNPs were identified where possible (e.g. tag SNPs for known GWAS hits). Otherwise, additional SNPs for low frequency coverage were added. The marker numbers given above reflect the final figures after array design.

### *C. Array Characteristics*

Out of 700,701 markers on the array, 354,399 are also present on the UK Biobank array.

Based on data collected during the array design process, predicted coverage of 1000 genomes CHB/CHS content was as follows:

| method | MAF | % $r^2$ ≥0.8 | Mean $r^2$ |
|---|---|---|---|
| imputation | ≥0.05 | 93.0 | 0.936 |
| imputation | ≥0.01 | 87.1 | 0.901 |
| imputation | 0.01-0.05 | 68.6 | 0.788 |
| pairwise | ≥0.05 | 68.9 | |
| pairwise | ≥0.01 | 67.5 | |
| pairwise | 0.01-0.05 | 62.9 | |

### China Kadoorie Biobank Array Revision

The overall strategy for revision of the CKB array design (summarised in **Figure S3**) was as follows:

- The overall performance of probesets on version 1 of the CKB array was assessed
- Probesets were identified for removal from the design on the basis of:
  - Redundancy (where a variant was interrogated by 2 probesets)
  - Assay failure or low quality
  - Low allele frequency (monomorphic in the first 100 plates of data and absent from other datasets), unless retained for other reasons
- Potential new content was identified including
  - Alternative assays for excluded probesets
  - Tag SNPs for excluded probesets
  - Novel content with putative functional effects
  - New GWAS hits
  - Additional content from collaborators
  - Improvements in/restoration of GWAS grid coverage

Included in this strategy was the use of the full sequencing dataset from the CONVERGE consortium[1], for both content identification and assessment of GWAS grid coverage.

#### *A. Array Version 1 Performance*

Two batches of 50 plates underwent standard QC; after exclusion of 5 plates that failed initial QC, a total of 8,995 datasets passed QC, including 98 duplicates. Genotyping of all probesets was carried out, and metrics were derived, using 0.98 as the call rate threshold.

#### *B. Identification of Probesets to be Removed*

Using the stated metrics output during genotype calling, SNPs/probesets were flagged for retention, exclusion, or review as follows:

- Retained: All HBV probesets [130 probesets];
- Excluded: Redundant probesets that were not the "preferred" probeset in either batch [57,223 probesets];
- Marked for Review (1): Probesets classed as "PolyHighResolution" or "NoMinorHom" in both batches and included in all "recommended" and "preferred" lists of probesets [585,939 probesets];
- Excluded: Probesets classed as any of "CallRateBelowThreshold", "OffTargetVariant" or "Other" in both batches [39,869 probesets];
- Marked for Review (2): Probesets classed as "MonoHighResolution" in one batch and either "PolyHighResolution" or "NoMinorHom" in the other, and included in all "recommended" and "preferred" lists of probesets [13,251 probesets];
- Marked for Review (3): Probesets classed as "MonoHighResolution" in both batches, and included in all "recommended" and "preferred" lists of probesets [30,628 probesets];
- Marked for Review (4): All hemizygous probesets from MT and chrY [1,162 probesets];

- Excluded: Probesets with a call rate <0.98 in at least one batch [8,570 probesets];
- Excluded: Remaining probesets that were not one of a pair of probesets but were not recommended in both batches [7,989 probesets];
- Excluded: Remaining probesets that in at least one batch were identified as "preferred" probesets but were nevertheless not recommended [5,964 probesets];
- Excluded: Remaining probesets that called one or both batches as "OffTargetVariant", "CallRateBelowThreshold" or "Other" [8,956 probesets];
- Excluded: Of each remaining pair of probesets, the probeset with the lower overall call rate [7,556 probesets], or if tied the lowest FLD [237 probesets], or if FLD comparison not possible the lowest HomRO [622 probesets];
- Marked for Review (5): Remaining probesets, recommended in one batch but not in the other, which were classed as "MonoHighResolution" in both batches [8,771 probesets];
- Marked for Review (6): Remaining probesets, recommended in one batch but not in the other, classed as "MonoHighResolution" in one batch and either "PolyHighResolution" or "NoMinorHom" in the other [2,246 probesets];
- Marked for Review (7): Remaining probesets, recommended in one batch but not in the other, classed as either "PolyHighResolution" or "NoMinorHom" in both batches [2,824 probesets].

Further review was carried out as follows:

1. Cluster statistics were checked for the reported FLD values. Probesets for which one or both batches had FLD<4.90 were excluded [21,528 probesets];
2. Cluster statistics were checked for the reported FLD and HomRO values. Probesets for which at least one batch had HomRO<0.4 were excluded (all of these were called as MonoHighResolution" in one batch and "PolyHighResolution" in the other) [91 probesets]. Probesets for which the non-monomorphic batch had FLD<4.90 were excluded [294 probesets];
3. These were reviewed together with probesets from (5).
   The frequencies of these SNPs in the CONVERGE dataset were checked. An appreciable number were found to have MAFs in CONVERGE that were sufficiently high that failure to identify any minor alleles was highly unlikely. Probesets corresponding to SNPs with a CONVERGE MAF>0.00158 (corresponding to $P<10^{-6}$; $P<10^{-3}$ even for probesets with 5 minor alleles in the first 2 batches of genotypes) were excluded [1456 probesets].
   Probesets for SNPs that were not found (at whatever frequency) in CONVERGE were excluded unless they were originally included on the array in one of the modules HLA/KIR (193), ApoE (493), Fingerprint (0), Neanderthal (887), LOF (2060), Ax-LOF (1535), GWAS hits (247), novel nonsense (39) [31,907 probesets].
4. Hemizygous SNPs were reanalysed with an updated version of SNPolisher, and treated as follows: (a) exclude duplicate probesets that were not preferred in either batch [65 MT probesets]; (b) exclude probesets for SNPs monomorphic in both batches, Taiwanese data and (for chrY) 1000 genomes CHB/CHS [660 chrY probesets, 62 MT probesets]; (c) examine cluster plots to select between remaining pairs of duplicate probesets, for similar-quality clustering selecting the probeset with higher call rate (or excluding them both) [17 MT probesets].
5. These were reviewed together with probesets from (3), see above.
6. Cluster statistics were checked for the reported FLD and HomRO values. Probesets for which at least one batch had HomRO<0.4 were excluded (all of these were called as

MonoHighResolution" in one batch and "PolyHighResolution" in the other) [16 probesets]. Probesets for which the non-monomorphic batch had FLD<4.90 were excluded [75 probesets];

7. Cluster statistics were checked for the reported FLD values. Probesets for which one or both batches had FLD<4.90 were excluded [282 probesets];

Additional probesets were excluded as follows:

Autosomal SNPs whose minor allele frequency gave an expected minor homozygote count of at least 5 (from review classes 1 and 7) were tested for Hardy-Weinberg disequilibrium. With Holm-Bonferroni multiple testing correction (5% family-wise error rate), probesets with $P<1.13x10^{-7}$ were excluded [2,734 probesets].

To further check SNPs with low MAF, the total minor allele count was extracted for those polymorphic SNPs still under consideration that were not "PolyHighResolution" in either batch (review classes 2 and 6). There was no obvious excess of SNPs with low minor allele count (1-3 minor alleles). Inspection of selected cluster plots did not indicate any problems.

### *C. Restoration of selected "monomorphic" SNPs*

The list of exclusions was checked for Fingerprint, CKB group, and GWAS hits and these were reviewed (2,410 in total).

7 Fingerprint SNPs restored

55 SNPs specified by the CKB group or collaborators were restored

For GWAS hits:

- Marginal call rate or QC failures were restored [776 variants];
- Variants with lower call rate were restored, but were not used in constructing the GWAS grid [159 variants];
- Monomorphic or near monomorphic probesets that failed initial QC were excluded.

After all exclusions and restorations were complete, 586,528 probesets were retained.

### *D. Selection of New Content*

Novel content was defined as follows (some variants were included for more than one reason):

#### *5. Novel functional content*

Coding variants (nonsynonymous, stop gain, etc.) were identified from CONVERGE. These were filtered to remove previously considered variants; this was achieved by excluding SNPs for which no 1000 genomes project frequency information was available. The remaining 72,332 variants were analysed by multiple functional prediction algorithms using TABLE_ANNOVAR. The results from these algorithms were combined to give an average score for whether a variant was deleterious – (sum of deleterious predictions)/(total number of predictions). Predictions were classed as deleterious as follows: SIFT – D=1; Polyphen_HDIV – D=1, P=0.5; LRT – D=1; Mutation_taster – A=1, D=1; Mutation_assessor – H=1, M=0.5; FATHMM – D=1; RadialSVM – D=1; LR_score – D=1.

Variants were selected for inclusion on the array if they had a score ≥0.5 derived from at least 3 algorithms, and had a variant-calling info score ≥0.1. As a result 9,619 variants were identified for addition to the array.

#### *6. New GWAS hits*

The NHGRI catalogue was downloaded on 28/4/15. There were 7,523 new entries since the list used for the original array design, of which 6,157 were for variants not previously included in the catalogue. Of these, 2,790 were associations at genome-wide significance ($P \leq 5x10^{-8}$), for 1,386 unique variants. 307 of these were already included on the array design, 2 were HLA haplotypes, and 49 were GxG interactions (for which a much higher P-value threshold would be appropriate).

As a result, 1,028 SNPs were identified for addition to the array.

#### *7. Additional content from collaborators*

Various external and internal collaborators supplied lists of variants, which were checked against the current array content. 292 additional variants were included.

Preliminary analysis of the HBV probes indicated they were successfully identifying HBV infection (strong association with HBV antigen test conducted at baseline). These results informed design of a further 24 HBV probes for inclusion on the array.

#### *8. Alternative assays/tags for important SNPs*

Key variants, specified by the CKB group and collaborators [15 variants] or which were GWAS hits [431 variants], that failed QC were marked as requiring alternative assays. Where possible, an assay from the opposite strand was designed, otherwise 'tag' SNPs ($r^2$>0.9) were selected from the Affymetrix library of validated assays.

### *E. Building of GWAS Grid*

The existing GWAS grid was patched and extended using similar procedures to those used during the original array design, with some modifications, as follows:

- Since completion of the original array design, low coverage sequence data from ~9,000 subjects from across China had become available from the CONVERGE Consortium[1]. These were used to update the allele frequency bins used to define the variant target list.
- The variant prioritisation criteria were updated to remove from consideration any variant already excluded from the array design
- Imputation aware variant selection was initially conducted simultaneously for all variants with MAF>0.01, and was halted once coverage of variants with MAF>0.05 reached 94.5% (an improvement on the previous 93%).
- Further greedy tagging of variants not covered by the 1000 genomes reference used both BGI and CONVERGE sequence data.
- Further imputation aware selection was conducted specifically targeting low-frequency variants (0.01≤MAF<0.05) and regions with poor coverage

In total a further 205,176 variants were added to the GWAS grid.

### *F. Array Characteristics*

Out of 803,030 markers on the array, 340,562 are also present on the UK Biobank array.

Predicted coverage of 1000 Genomes CHB/CHS content is as follows:

| **method** | **MAF** | **% $r^2$ ≥0.8** | **Mean $r^2$** |
|---|---|---|---|
| imputation | ≥0.05 | 93.3 | 0.942 |
| imputation | ≥0.01 | 85.3 | 0.900 |
| imputation | 0.01-0.05 | 63.1 | 0.766 |
| pairwise | ≥0.05 | 73.9 | |
| pairwise | ≥0.01 | 67.5 | |
| pairwise | 0.01-0.05 | 62.3 | |

Note that there were some changes in the target sets, particularly for the low frequency bin, so these results are not directly comparable to those for the original array design.

### Supplementary Methods

*DNA extraction and SNP genotyping*

For DNA extraction at BGI, Shenzhen, using KingFisher™ Blood DNA Kit and KingFisher™ Flex 24 Magnetic Particle Processors (Thermo Scientific), buffy coat sample tubes were barcode scanned, and up to 800µL was manually pipetted into the extraction tube at the positions specified by a bespoke sample-tracking IT system. Extracted DNA was transferred using a Freedom EVO® (Tecan) fluid handling system, up to 200µL to each of two sets of 96-tube racks of 2D-barcoded cryovials (Fluidx, Azenta Life Sciences), and 12µL to a 96-well microtitre plate. DNA concentration and quality was recorded using a NanoDrop Microvolume Spectrophotometer (Thermo Scientific). Tubes were frozen and shipped on dry ice to the CKB sample storage facility in Beijing, for long-term storage at –70C. All sample movements were recorded by the sample-tracking IT system.

SNP genotyping was performed in 96-well microtitre plates, and used up to 5µL DNA from the microtitre plate. Each genotype plate included positive and negative controls at fixed positions, and 2 pairs of duplicate samples at unique combinations of plate positions. GoldenGate 384-plex genotyping was performed for a total of 1,040 plates according to GoldenGate Genotyping Assay Manual Protocols[2], with beadchip imaging using an iScan System (Illumina). The 384 SNP panel was revised after genotyping of the first 100 plates (9200 unique samples), and again after the second 100 plates. Genotyping calling was performed in 4 batches using GenomeStudio software, with initial QC based on automated clustering. All negative controls had a SNP call rate of 80% or less (mean = 34%). 15 plates were flagged for inspection due to an initial positive control call rate <95%, but no failures of genotyping were identified; the remaining positive controls had mean call rate of 98.6%. A further 12 plates were flagged for inspection due to 1 or both of 12 pairs of duplicates being among 709 samples excluded with call rate <90%, but again no failures of genotyping were identified; for 2,063 remaining pairs of duplicates, genotyping concordance was between 98.66% and 100% (mean 99.98%).

Following this initial QC, and again after final sample QC, SNPs were reclustered, and within each batch SNPs with GenTrain score <0.7 were inspected manually, and manually reclustered or excluded as appropriate. Across the 3 SNP panels, 30 SNP assays failed genotyping within that panel (either due to gross genotyping failure or call rate <95%), and a further 42 SNP assays failed for a subset of the 4 'plexes'. Two SNPs displayed Hardy-Weinberg disequilibrium due to assay interference by nearby SNVs or indels. 15 SNPs displayed potential batch effects, identifying genotype clustering errors that were adjusted manually.

Following SNP QC, an additional 1,518 unique samples (2,217 in total) were excluded on the basis of a SNP call rate <98%. For 118 samples, mismatches of reported gender were identified with inferred sex based on 3 sex-informative SNPs from chrXY pseudoautosomal regions, and a further 136 samples from blocks with multiple sex-mismatches or with other potential sample linkage errors were also excluded. One sample with excess heterozygosity (F-statistic >5 SDs above the mean) was excluded.

*Genome-wide genotyping*

For genotyping using the first version of the CKB array, samples were selected for genotyping as part of nested case-control or case-cohort study designs. Incident cardiovascular disease cases were selected according to available disease follow-up at time of sample selection (August 2014) from amongst those with extracted DNA and no self-reported prior cardiovascular disease history, as follows: (a) all cases of intracerebral haemorrhage (ICH – CD-10: I61, I69.1) where this was the first stroke event, including additional samples selected for prioritised DNA extraction and one case originally incorrectly recorded as an ischaemic stroke (IS); (b) all available cases of subarachnoid haemorrhage (SAH – ICD-10: I60, I69.0) where this was the first stroke event; (c) 5,662 cases of IS (ICD-10: I63, I69.3) occurring prior to 1 January 2014 at age ≤71 years where this was the first stroke event; (d) 1,008 incident cases of myocardial infarction (MI – ICD-10: I21-I23); and (e) all available cases of death with ischaemic heart disease as underlying cause (fatal IHD – ICD-10: I21-I25). Pairs of controls with no cardiovascular disease events or self-report were identified for each ICH case, matched to sex, recruitment region, and year of birth. For respiratory disease, 5,358 participants were selected with at least one event of hospitalisation with chronic obstructive pulmonary disease (COPD – ICD-10: J41-J44); as controls, 4,766 participants were randomly selected from amongst those who attended the second resurvey. For genotyping using the second version of the CKB array, selection was on the basis of complete boxes of DNA samples, prioritising those boxes that contained samples from participants originally recruited in clinics at which the second resurvey was conducted. These samples were supplemented with additional cases of ICH, SAH, MI, and fatal IHD that occurred subsequent to initial sample selection.

DNA samples were retrieved from storage at –70C, either as complete boxes of 96 samples or (for nested case-control samples) individually selected and transferred to new boxes, and were shipped on dry ice to BGI, Shenzhen. DNA concentration was checked using a NanoDrop Microvolume Spectrophotometer (Thermo Scientific), and a Microlab STAR liquid handling system (Hamilton) used for transfer of sub-aliquots to new racks of 96 Fluidx cryovials and dilution with TE buffer to 80 ng/µL; the equivalent measured concentration of a subset of samples measured using Qubit DNA quantification (ThermoFisher) was 50ng/µL. Diluted DNA was plated onto 96-well microtitre plates, with samples from a minimum of 3 boxes distributed across a single plate (a 1:1 mix of cases and controls for nested case-control samples). Samples with low DNA concentration were plated separately for genotyping with a modified first stage of the protocol, using a larger volume of DNA in place of TE buffer. Samples at position H12 were replaced with a duplicate sample from position D1 on the previous plate, thereby providing checks of genotyping quality and sample tracking. Genotyping was performed with manual target preparation according to Affymetrix protocols and automated plate processing and imaging using GeneTitan® Instruments[3]. Raw genotyping data were exported from China to the Oxford CKB International Coordinating Centre under Data Export Approvals 2014-13 and 2015-39 from the Office of Chinese Human Genetic Resource Administration.

Genotyping quality control (QC) and calling was conducted, for each array version separately, according to Affymetrix Best Practice workflow[4] using the Axiom Analysis Suite (Affymetrix) with default settings. Initial QC was performed on samples genotyped on batches of 50 plates. Genotyping was carried out for a preselected set of ~20k "high performance" SNPs, using the 'Sample QC' option, to give initial quality metrics. These were used to identify samples and plates to be excluded from subsequent steps, on the basis of sample DQC<0.82; sample QC call rate <97%; or plates with mean call rate for remaining samples <98.5%. Plates with sample pass rate <95% were flagged for inspection, and were excluded if there was evidence of a general failure of genotyping (e.g. large sections of the plate have failed), or if sample call rate was systematically low relative to sample DQC (rather than having a large number of failing samples due to e.g. a group of samples with poor quality DNA). Some plate failures identified array manufacturing defects; genotyping of these plates was repeated using a new array. Samples passing initial QC were then processed and co-clustered, again in batches of 50 plates, to derive genotypes and further quality metrics.

*Variant QC*

Within each batch, probesets were "failed" if they were classified as "OTV" (off target variation), "CallRateBelowThreshold" (using the default threshold 95%), or "Other", and genotypes for non-failed probesets; all further QC was performed using PLINK v1.9 and/or v2.0[5]. Within each batch, probesets were assessed for the presence of plate effects: logistic regressions were conducted to test each individual plate within a batch for significant deviations in genotype calling: each plate in turn was treated as "case" status with all other plates in the batch as controls, with recruitment centre as covariate; probesets were failed according to criteria determined empirically through manual review of cluster plots to identify clustering failures – any plate effect with $P<10^{-10}$, >3 instances of plate effect $P<10^{-4}$, or any plate effect $P<10^{-8}$ and clustering metrics FLD<8, HetSO<0.68, and HomRO<3.7; in addition, for probesets with any plate effect $P<2x10^{-5}$, cluster plots were manually reviewed, and appreciable clustering failures (e.g. poor cluster separation) were "failed".

Probesets passing this initial QC were combined into a single dataset, and a preliminary round of sample QC was performed (see below). An LD-pruned set of autosomal probesets (PLINK option `--indep-pairwise 50 5 0.1`) was then used to identify an unrelated subset of samples (PLINK `--rel-cutoff 0.025`). These were used to test for significant deviations in genotype calling between batches: logistic regressions were performed treating each batch in turn as "case" status with all other batches as controls, again with recruitment centre as covariate; probesets were failed entirely, across *all* batches, again according to criteria determined empirically through manual review of cluster plots to identify clustering failures: probesets with any batch effect with $P<10^{-10}$, >2 or >7 with $P<10^{-4}$, for array versions 1 and 2 respectively. Clustering was manually checked for remaining probesets with a batch effect with $P<10^{-3}$ and were scored as either "Pass", "Batch Fail" (fail in one batch only), or "Fail". Probesets failing in >10% of batches (i.e. any batch for array version 1, >1 batch for version 2), or with call rates <98% (in passed batches) were excluded entirely from the dataset for that array version.

Probesets were then tested for deviation from Hardy-Weinberg equilibrium (HWE): tests were performed in each recruitment region separately (PLINK `--hardy midp`) using unrelated individuals (women only for chrX variants), and probesets with a HWE $P<10^{-6}$ (10 degree of freedom sum-of-Chi-squared test) were excluded. In addition, variants with a minor allele frequency (MAF) >0.2 different from that in the 3 Chinese populations from the 1000 Genome Project Phase 3 reference[6] were excluded, and one pair of duplicate probesets assaying the same variant (that with the lower call rate) was removed.

Performance of version 2 of the array was assessed using data for 192 samples (152 Chinese, 40 European) from the European Vasculitis Genetics Consortium[7] which had been genotyped with both the CKB_2 and UK Biobank Axiom® arrays. For 331,838 probesets passing QC on both arrays, concordance between the two arrays was assessed (PLINK `--merge-mode 7`): 99.5% of genotypes were non-missing for the data from both arrays, with a concordance of 99.80%.

*Sample QC*

Primary sample QC was conducted for each assay version separately. Samples were excluded which had genotyping call rate <0.95, or high/low heterozygosity determined as follows: sample heterozygosity was assessed for autosomal variants with MAF>0.01 (PLINK `–het` followed by calculation of heterozygosity as 1–HOM/NMISS), mean and SD was determined for samples from each recruitment region (Note: there was a clear North-South gradient in heterozygosity, with a range of values >1 SD), and samples with a region-specific Z score >+3 were excluded; total runs of homozygosity were determined for each sample (PLINK `--homozyg-kb 1000`), and 3 samples with a region-specific Z score <–3 *and* a Z-score <2 for total runs of homozygosity were excluded (**Supplementary Figure S7**).

Samples from individuals with appreciable non-Chinese ancestry were identified by projecting CKB participants onto principal component analysis of the 1000 Genomes Project Phase 3 reference populations[6] using an LD-pruned set of 104,866 variants with MAF >0.01, passing QC for both CKB array versions, and excluding major regions of long-range LD[8] (PLINK `--pca --within --pca-clusters`) (**Supplementary Figure S13**). A total of 4 individuals were excluded who had a PC value >10 SDs from the CKB-wide mean for at least one of the first 10 PCs.

Initial checks of computed sex with that reported in participant data (PLINK `--check-sex`) identified multiple clusters of sex mismatches, indicating systematic linkage errors. All such clusters of mismatches were tracked back through all steps of sample handling, and the majority could be unambiguously traced to specific sample-handling errors (e.g. 180° rotation of boxes of DNA samples), such that correcting such sample-linkage errors removed all instances of sex mismatch in a cluster without leading to new ones. For clusters that remained uncorrected, all samples in the affected block of samples, irrespective of sex mismatch, were marked for exclusion from the dataset. Other individual sex-mismatched samples were also excluded.

For more detailed checks for sex mismatch, the chrY/chrX probe intensity ratio (parameter cn-probe-chrXY-ratio_gender_ratio output to file AxiomGT1.report.txt during genotyping) was plotted against the chromosome X heterozygosity F-statistics (from PLINK `--check-sex`), grouping genetically male and female samples into distinct clusters and clearly identifying sex mismatches (**Supplementary Figure S14A**). In addition, groups of samples were observed representing potential chromosome XY aneuploidies, including Klinefelter Syndrome and non-Klinefelter XXY, and XO (Turner Syndrome) or XXX, and phenotypic males with appreciable chrX heterozygosity and lower than average chrY/chrX probe ratio. These latter individuals may include individuals with partial chrX translocations, but for most of them the heterozygous markers were distributed along the length of chrX. The samples corresponding to phenotypically male participants were clearly identifiable and were excluded without further investigation. To more robustly identify aneuploid female samples, probe intensity data was extracted using Affymetrix Axiom® CNV Tools software[9], and 363 samples were identified whose mean probe intensity (LRR) on chrX was >3 SDs from the mean; for these, probe heterozygosity (BAF) was visualised across chromosome X, enabling identification of 6 Turner, 10 Turner mosaic, and 36 XXX individuals, either with no chrX heterozygosity (Turner) or with BAF values for heterozygous states consistently different from 0.5 (**Supplementary Figure S14B**); other aneuploidies were also identified including a partial deletion of the p-arm of chrX and a complex rearrangement with both a partial q-deletion and partial p-duplication. All these individuals were marked for exclusion.

The datasets for the two array versions were merged into a single dataset, and genetically identical samples with PI_HAT of ~1.0 were identified by testing pairwise relatedness using an LD-pruned and thinned set of 10k autosomal SNPs with MAF>0.05 (PLINK `--thin-count 10000 –make-rel`). All expected duplicate pairs (including a small number of samples genotyped twice in error) were identified, confirming correct genotyping plate layout and order. All unexpected duplicate pairs were resolved as due either to repeat samples from the ~2,000 individuals known to have attended the baseline survey twice, or to pairs of individuals whose personal data at recruitment (e.g. recruitment location, date of birth) supported their assignment as putative monozygotic twins. For each pair of duplicate samples, the dataset with the lower call rate was excluded.

*Imputation*

Further QC was performed prior to imputation. Variants were excluded which were at multiallelic locations, or which had alleles in CKB that did not match those in the 1000 Genomes Project Phase 3 reference[6] (October 2014 release); where indicated, strand-flips were performed to match the reference. In addition, to avoid potential phasing or imputation batch effects, variants were excluded if they failed QC in any genotyping batch. For imputation across the whole cohort, only variants present and passing QC in all batches on both arrays were included in the merged dataset. Imputation included samples excluded from the main dataset on the basis of sex mismatch, linkage errors, or chromosome XY aneuploidy (autosomal imputation only). Phasing was performed for entire chromosomes using SHAPEIT3 r882[10] with default parameters, except for chromosome X (SHAPEIT2 v2.17[11] with the -X option, and with pseudoautosomal regions excluded). For imputation, the reference panel was filtered to exclude variants with MAF=0 in the 5 East Asian populations, leaving 24759908 variants, and was split into 713 chunks, with length ranges from 330Kbp to 5264Kbp, mean 3948Kbp. Imputation was conducted in 20 batches of samples, using IMPUTE4 v4.r265[12] for autosomes and IMPUTE2 v2.3.2[13] for chromosome X, with buffer regions of 500Kbp. Subsequent to imputation, checks for batch and array-version effects were conducted by testing for association using BOLT-LMM v2.3.1[14] with individual batches or array version as binary variables; 3867 variants displaying significant batch effects ($P<5x10^{-8}$) were excluded from the imputed dataset.

*Relatedness*

Relatedness between individuals was assessed using an LD-pruned (PLINK `--indep-pairwise 50 5 0.1`) set of 122,675 variants with MAF>0.01. On the assumption that near relatives were not present in different recruitment regions, identity-by-descent was determined for all pairs of individuals within each region (PLINK `--genome gz`). First-degree (parent-child, siblings including twins) and second-degree (grandparent-grandchild, uncle/aunt-niece/nephew) relatives were identified as pairs of individuals with PI-HAT>0.375 and PI-HAT>0.1875, respectively, thresholds which very clearly resolve these relationships; from the first-degree relatives, parent-child pairs were identified as those with Z0 <0.05 and Z1 >0.5, with the parent identified as the older of the pair. Each pair of siblings was checked for the number of recorded first-degree relatives in the dataset, and the family structures of mismatches were investigated, leading to identification of one instance of 2 sets of putative three-quarter siblings.

*Principal component analysis*

PCA was conducted using FlashPCA v2.1[15] after LD pruning and exclusion of regions of long-range LD which, if not excluded or otherwise accounted for, can interfere with PCA potentially leading to erroneous conclusions about population structure, or to erroneous genetic association signals. Initial PCA excluded previously-identified regions of long-range LD[8] and used a heavily LD-pruned set of SNPs but, as previously noted for UK Biobank[16], visualisation of variant weights revealed that multiple PCs were nevertheless affected by disproportionate contributions from particular regions of the genome, likely reflecting further regions of long-range LD present in the Chinese population (**Supplementary Figure 15**).

We conducted a systematic iterative search to identify and remove regions of long range LD that influenced PCA in this way. Initially, PCA was conducted using an LD-pruned (PLINK `--indep-pairwise 50 5 0.1`) set of 122,675 variants with MAF>0.01, for 76,719 unrelated CKB participants. Leading PCs were checked for the presence of long range LD regions, pairs of identified regions closer than 1Mbp were merged into single extended regions, variants within those regions were excluded, and the PCA was repeated. This process was continued until no long range LD regions were identified in any of the leading 11 PCs informative for CKB population structure, nor in the 12$^{th}$ (not informative) PC. Long range LD regions were identified using a hidden Markov model: presence within/outside a long range LD region was the hidden state; transition between states was in proportion to EAS recombination rates (downloaded from SNiPA[17]); and emission was the posterior probability of being in a long range LD region given the square of the Z-score for the variant loadings for that PC. Variants were identified as within a long range if they had a posterior marginal probability >0.5. A total of 223 regions were identified (**Supplementary Table S9**) leaving a set of 171,236 variants for use in PCA.

To identify PCs informative for population structure of the full CKB dataset, models were constructed predicting individuals' recruitment region in which the top PCs were progressively added to the model, using multinom() from R package 'nnet'[18], and Bayes Information Criterion (BIC) was derived using the R BIC() function; the same approach was used to identifying PCs informative for self-reported Han status in Liuzhou ('Han', 'mixed', 'non-Han'). To identify PCs informative for population structure within each recruitment region, BIC was similarly determined for linear models (using the R lm() function) predicting the latitude and longitude of the study clinics at which participants were recruited. Informative PCs were those that reduced BIC when added to the model.

*Analysis subsets*

Subsets of the full genotyped dataset were derived for different analysis approaches (**Supplementary Table S8**). For region-stratified analyses, samples with non-local ancestry were excluded; these were identified as outliers for one or more of the informative PCs for that region, on the basis of a robust Mahalanobis distance (from the R mahalanobis() function) of >3 SDs. For analyses requiring unrelated individuals, these were defined using 122,675 LD-pruned variants with MAF>0.01, as above, but applying PLINK `--king-cutoff 0.05` which generally gives a larger set of unrelated individuals than `--rel-cutoff`.

Construction of a subset of genotyped individuals that was largely representative of the overall CKB cohort was based on the fact that the majority of genotyped samples were not selected individually but as complete boxes of DNA samples. These boxes of DNA were prioritised for genotyping solely according to the number of samples they contained that were from participants recruited at study clinics subsequently used for the second resurvey; these clinics had themselves been selected to be population-representative. The procedures for sample collection and DNA extraction meant that each box of DNA included a mixture of samples from at least two randomly-selected boxes of buffy coat samples. Therefore, samples in boxes of DNA were either from individuals invited to the second resurvey and therefore largely representative of the overall CKB cohort, or were random collections of samples from other recruitment locations.

An initial attempt to construct a cohort-representative subset used samples from those boxes with at least 70% of samples genotyped (irrespective of QC), but this was found to be *depleted* for certain ascertained disease cases; this was due to the early prioritisation of a proportion of ICH, SAH, and fatal IHD cases, which led to the transfer of these samples to different storage locations prior to DNA extraction. Therefore, the CKB-representative subset was instead based on the boxes in which blood samples were originally stored immediately after collection and processing at time of recruitment, and used samples originating from boxes of buffy coat with ≥40% of samples selected for genotyping. This gave a set of 77,176 participants which were representative of the overall CKB cohort, in which over-representation of the ascertained diseases was eliminated.
