## Supplementary Figures for "Genotyping and population structure of the China Kadoorie Biobank"

**Figure S1. CKB Axiom® array design overview.** The figure summarises the data sources used for array design and the filtering, QC, and variants selection procedures applied.

**Figure S2. Design of the CKB Axiom® genotyping array.** The figure illustrates the different categories of content on the initial CKB array. Numbers indicate the approximate counts of variants in each category. Some variants fall into more than one category.

**Figure S3. CKB Axiom® array design revision overview.** The figure summarises the procedures used to update the array design.

**Figure S4. Allele frequency of genotyped variants in CKB regions.** Allele frequency of QCed variants on array v2 in each CKB region, compared with the corresponding allele in the East Asian subset of the 1000 genomes Phase 3 reference.

**Figure S5. Imputation quality for each CKB array version.** The distribution is shown for imputation INFO score for variants in 4 MAF ranges, for the results of imputation using genotyping data from each CKB array version separately.

**Figure S6. Patterns of relatedness in CKB regions.** The histograms show for each CKB region the distributions of the relatedness between all possible pairs of genotyped individuals.

**Figure S7. Quality control for heterozygosity and homozygosity.** Overall heterozygosity and total runs of homozygosity were determined for each genotyping dataset. Blue symbols denote samples with low heterozygosity that is accounted for by extended runs of homozygosity. Red symbols denote samples whose heterozygosity is not accounted for by runs of homozygosity and which were excluded from the analysis dataset.

**Figure S8. Principal component analysis of CKB.** The results of PCA of the full CKB genotyped dataset are shown for pairwise plots of all PCs that were informative for CKB recruitment region. Data points are colour coded according to the region from which that participant was recruited.

**Figure S9. Population structure in CKB regions as informed by whole cohort PCA**. Local maps are shown for each recruitment region, showing the geolocation of the individual recruitment clinics, colour coded according to latitude and longitude; the size of the symbol is proportional to the number of genotyped individuals from that clinic. Corresponding PCA plots show the first two principal components from PCA of the full CKB cohort, colour coded according to their recruitment clinic. Top 2 rows ¬– urban regions; bottom 2 rows – rural regions.

**Figure S10. Identification of informative principal components for CKB regions.** Models predicting the latitude (blue) and longitude (red) of participants' recruitment clinic were constructed by progressively adding PCs, and Bayes Information Criterion was determined. Broken lines – PCA of the entire CKB cohort; solid lines – PCA of each region separately.

**Figure S11. Population structure in Liuzhou region.** Participants recruited in Liuzhou who attended the second resurvey are plotted according to PCA from the entire CKB cohort (top) or Liuzhou only (bottom). Green – self-reported Han ancestry; orange – mixed ancestry; red – non-Han ancestry. Plots (right) show the Bayes Information Criterion for models predicting Han status using increasing numbers of PCs.

**Figure S12. Population diversity in CKB.** Population differences as measured by Fst were derived, and trees were constructed to illustrate the relationships between them the populations shown. (A) Phylogenetic tree derived using the full unrelated CKB dataset, except for Liuzhou (RC46) for which only second resurvey participants were included. (b) Neighbour-joining tree constructed using 100 unrelated individuals from each population. RC12 – Qingdao; RC16 – Harbin; RC26 – Haikou; RC36 – Suzhou; RC46 – Liuzhou; RC52 – Sichuan; RC58 – Gansu; RC68 – Henan; RC78 – Zhejiang; RC88 – Hunan; CHB, CHS, JPT, CDX, KHV – East Asian 1000 Genomes populations.

**Figure S13. PCA projection onto 1000 Genomes.** PCA was conducted for the 1000 Genomes Phase 3 populations, and CKB participants were projected onto the resulting PCs. Top – 1000 Genomes populations; bottom – with CKB participants (black) included.

**Figure S14. Identification of sex mismatches and chromosome XY aneuploidies.** (A) plot showing relationship between chromosome X homozygosity and chromosome XY probe ratio. Sex-mismatched samples are visible within the main clusters of females (red) and males (blue). Open symbols denote samples identified as potential aneuploidies. (B) Plots across chromosome X of the BAF parameter which reflects the proportion of signal on the genotyping array coming from the two possible alleles at each site. 3 classes of aneuploidy are illustrated, the red marks highlighting systematic deviations from the expected 3 possible genotypes.

**Figure S15. Distortion of PCA by regions of long range LD.** Plots show individual variant loadings (Z-squared), for each of the first 12 PCs from PCA of the full CKB cohort, that results if regions of long range LD are not fully excluded.

**Figure S16. Identification of informative principal components.** Models predicting participant recruitment region were constructed by progressively adding PCs from PCA of the full CKB cohort, and Bayes Information Criterion was determined.

Supplementary Figure S1

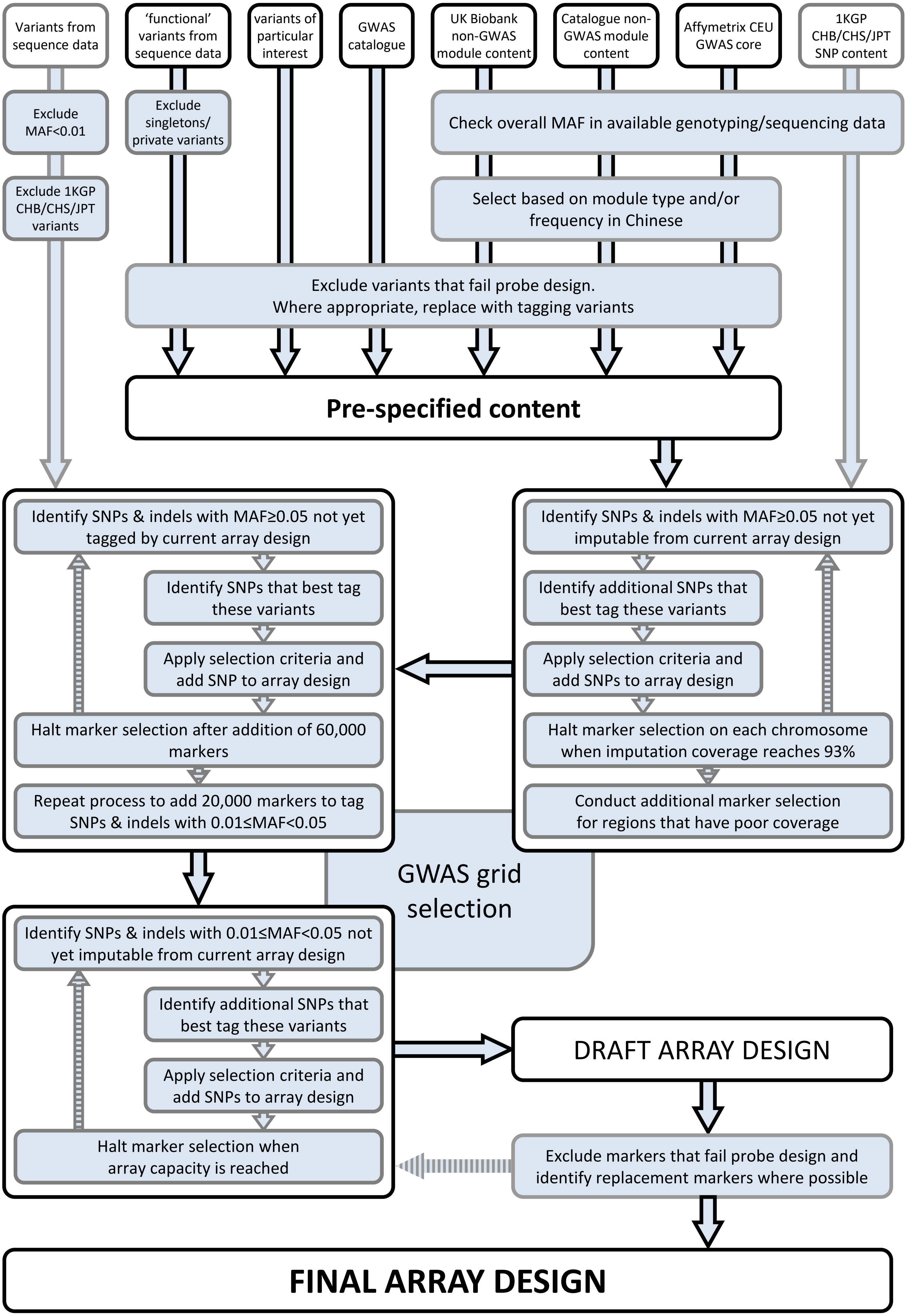

Supplementary Figure S2

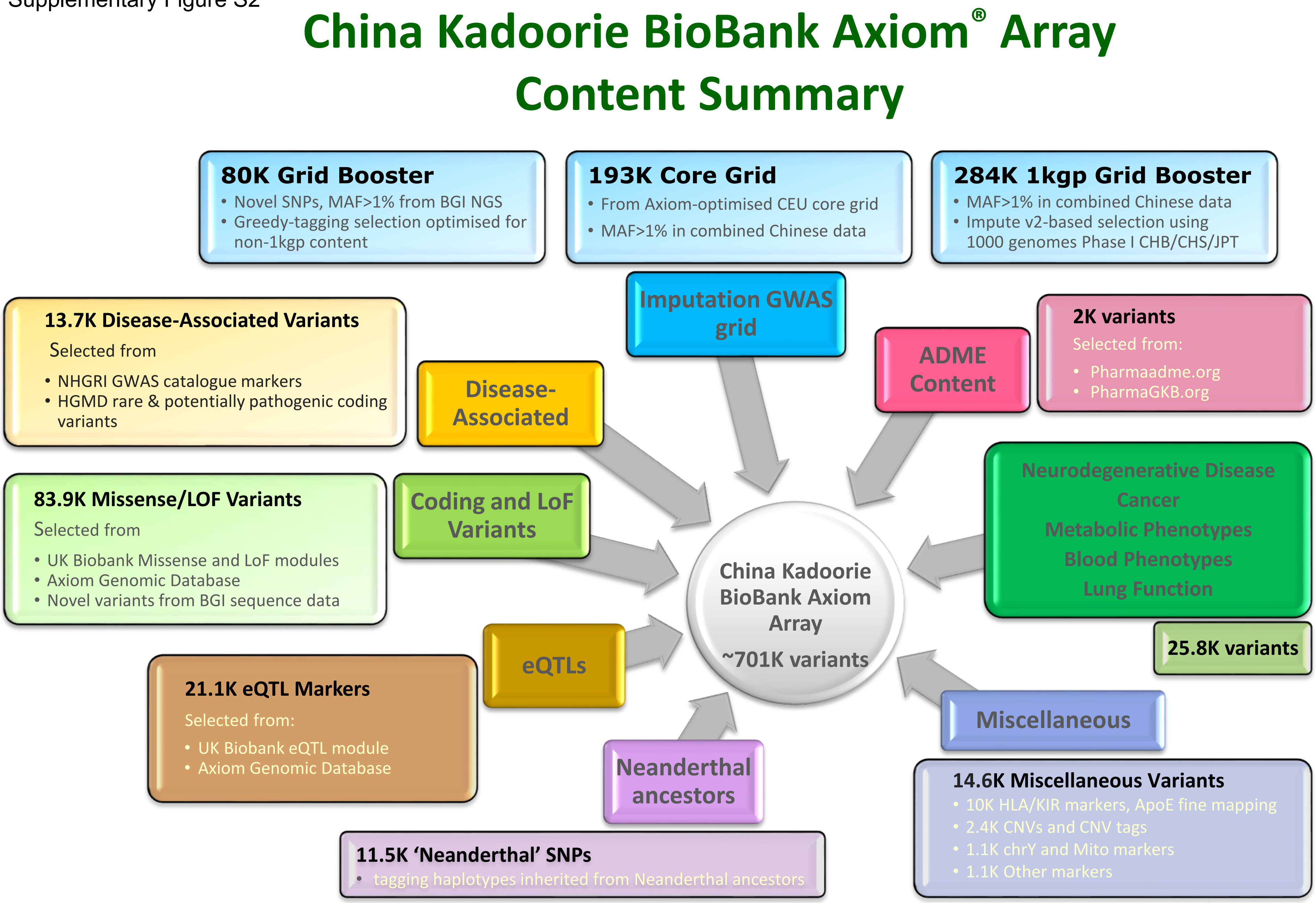

Supplementary Figure S3

**Review results from first 100 plates**

| | |
|---|---|
| Poly High Resolution | Review, default=retain |
| No Minor Homozygote | Review, default=retain |
| Mono High Resolution | Review and check frequency in CONVERGE |
| Hemizygous | Review |
| Off Target Variant | Review, default=exclude |
| Call Rate Below Threshold | Review, default=exclude |
| Other | Review, default=exclude |
| Viral probes | Review, default=retain |

Identify unneeded 2nd probesets
Check call rate for all retained SNPs

**Remove from design**

- Failed and low-quality assays
- Non-preferred probesets
- Monomorphic variants (except for selected variants e.g. those confirmed at acceptable frequencies in CONVERGE)
- Viral probes?

**Add to design**

- Modified assays for retained SNPs
- Tag SNPs for important failed assays
- Novel content from CONVERGE
- New GWAS hits
- Additional content from collaborators

**Augment GWAS grid**

- Restore gaps caused by SNP removal
- Supplement low freq. grid

Supplementary Figure S4

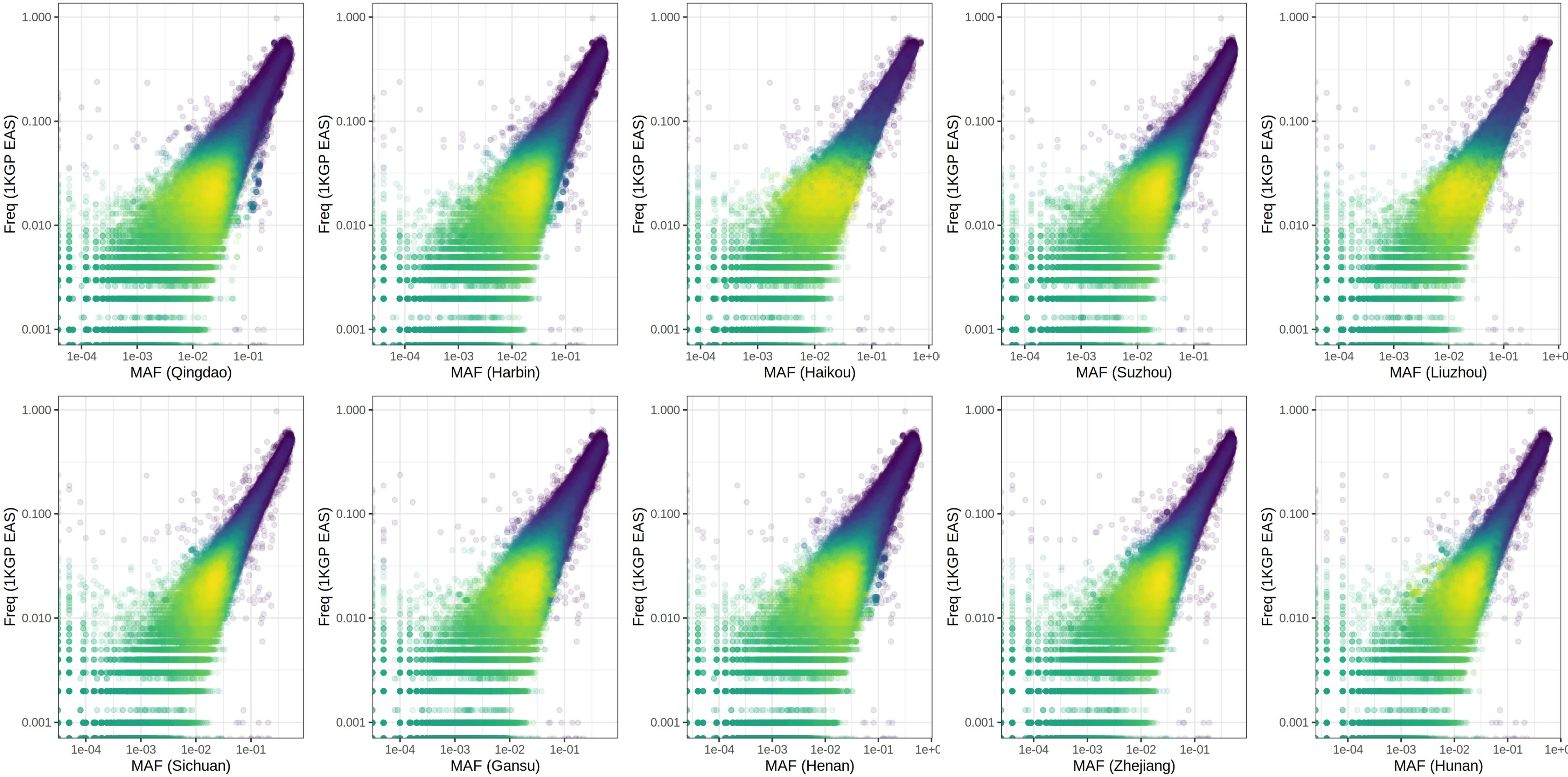

Supplementary Figure S5

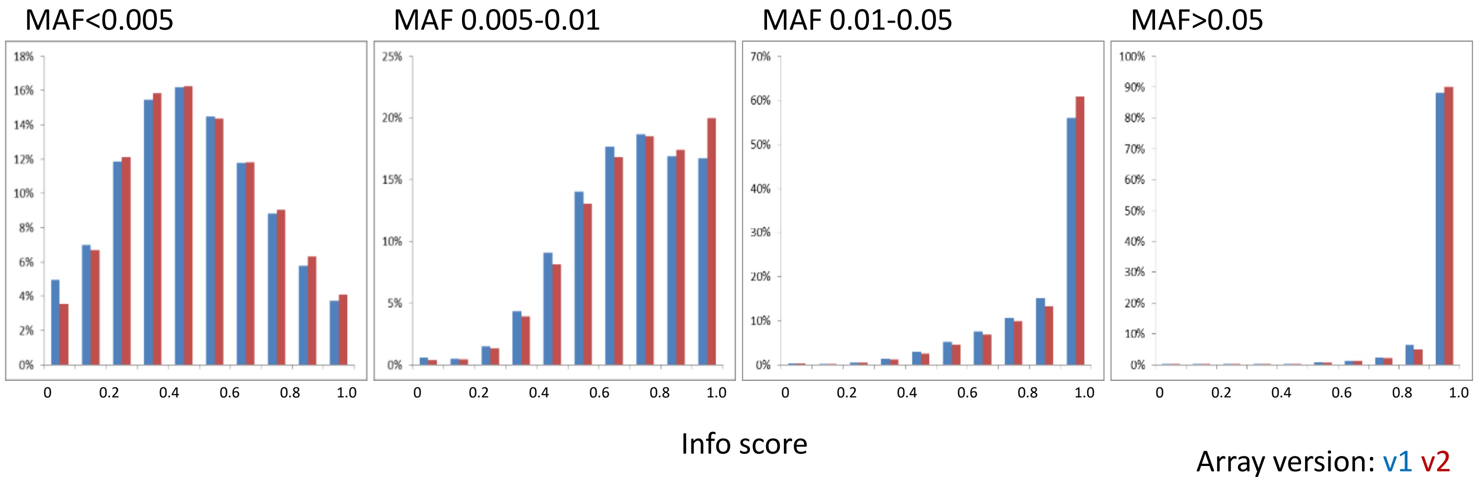

Supplementary Figure S6

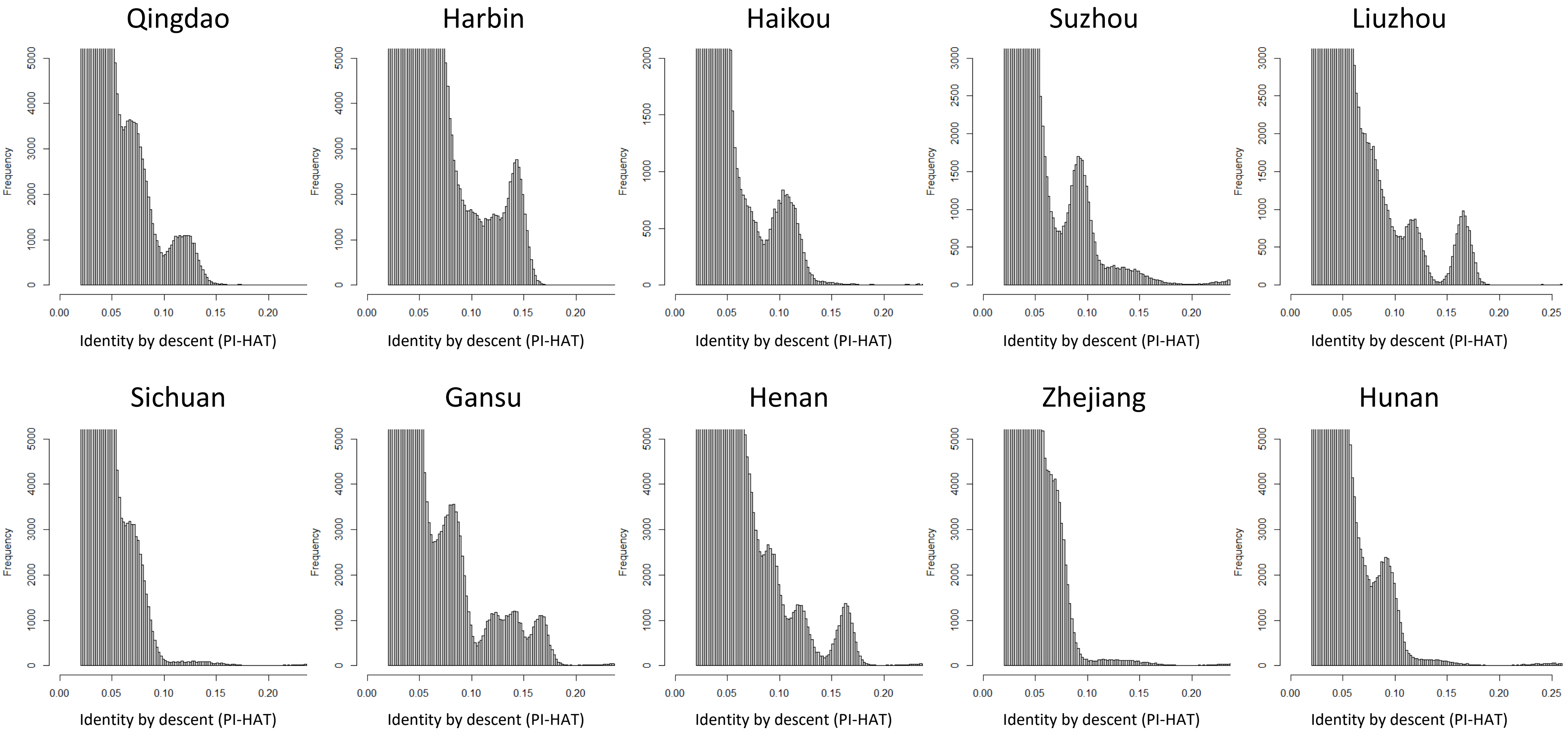

Supplementary Figure S7

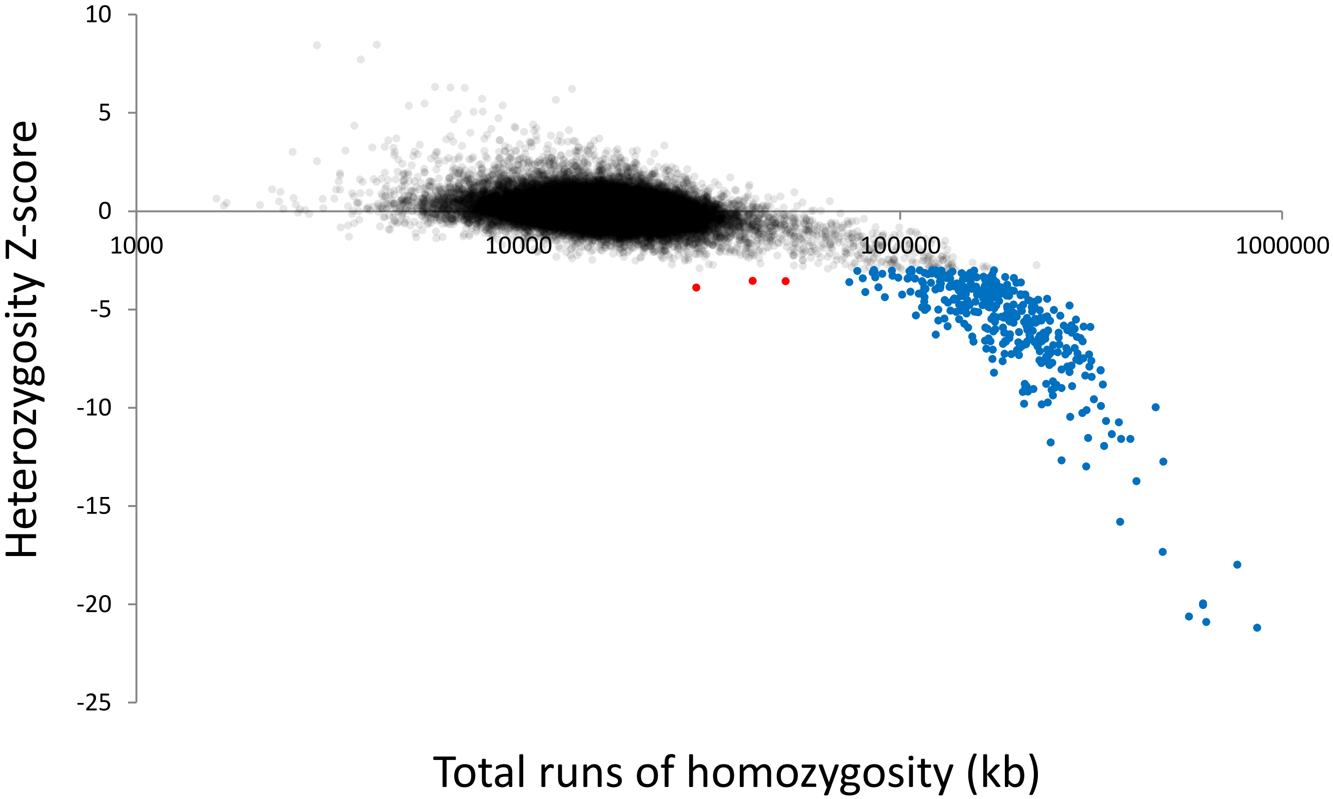

Supplementary Figure S8

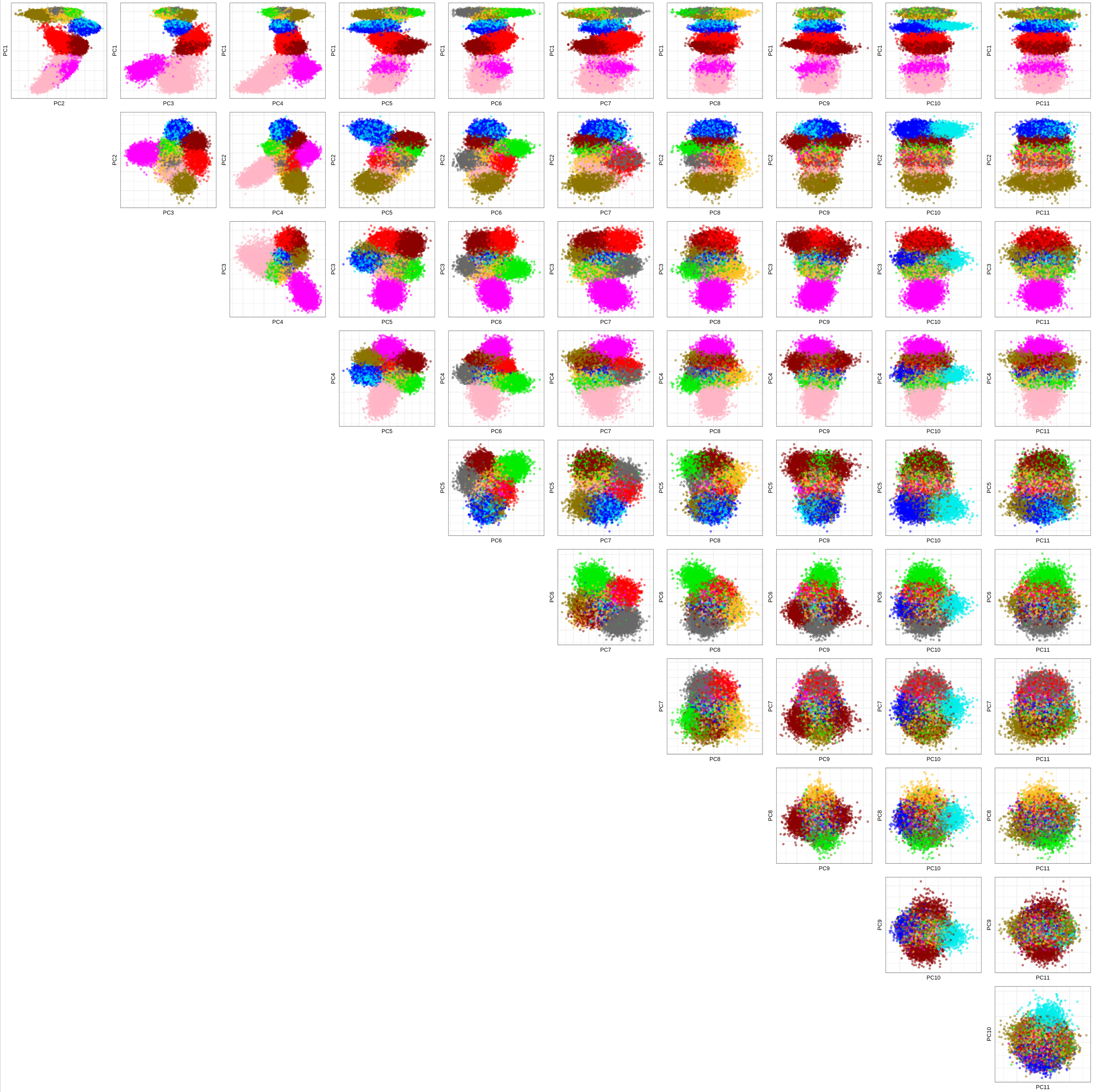

Supplementary Figure S9

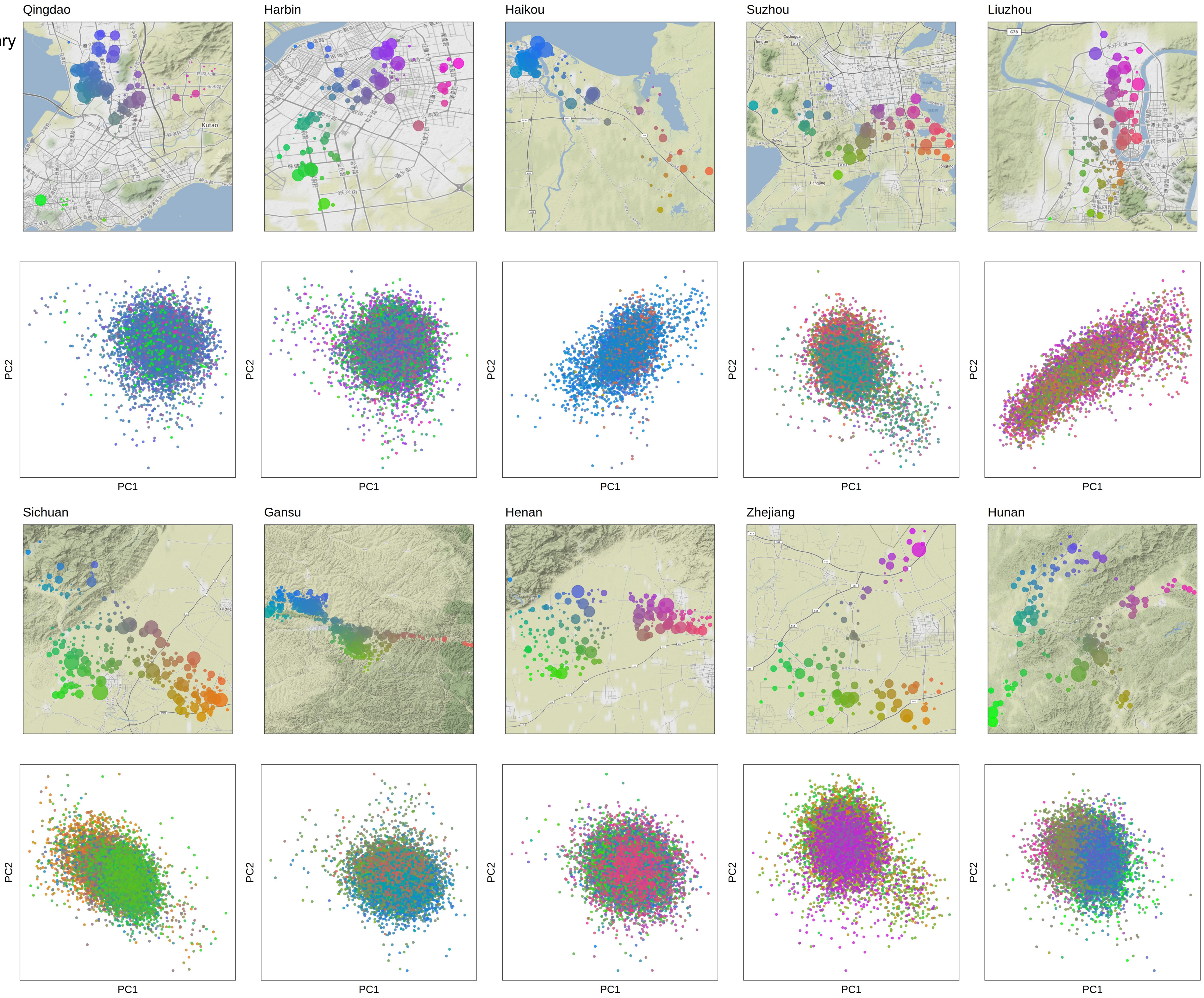

Supplementary Figure S10

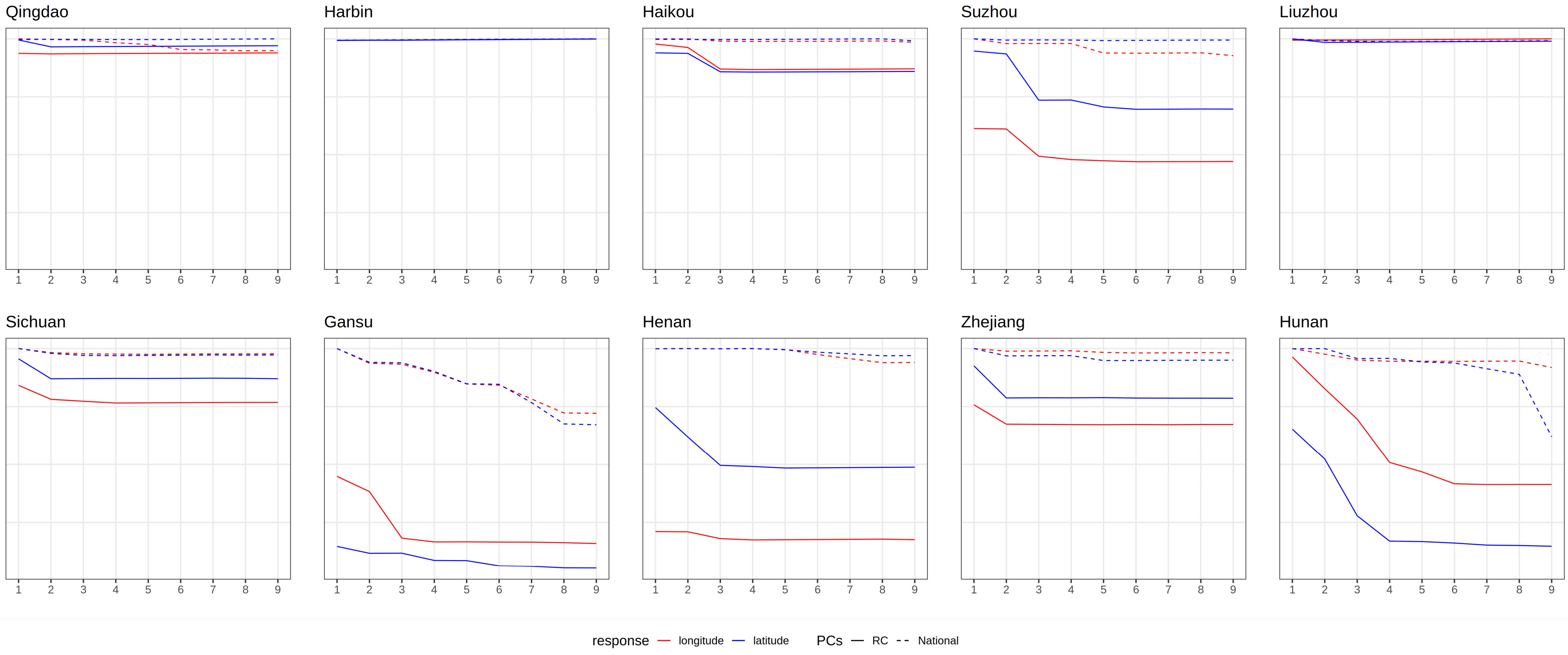

Supplementary Figure S11

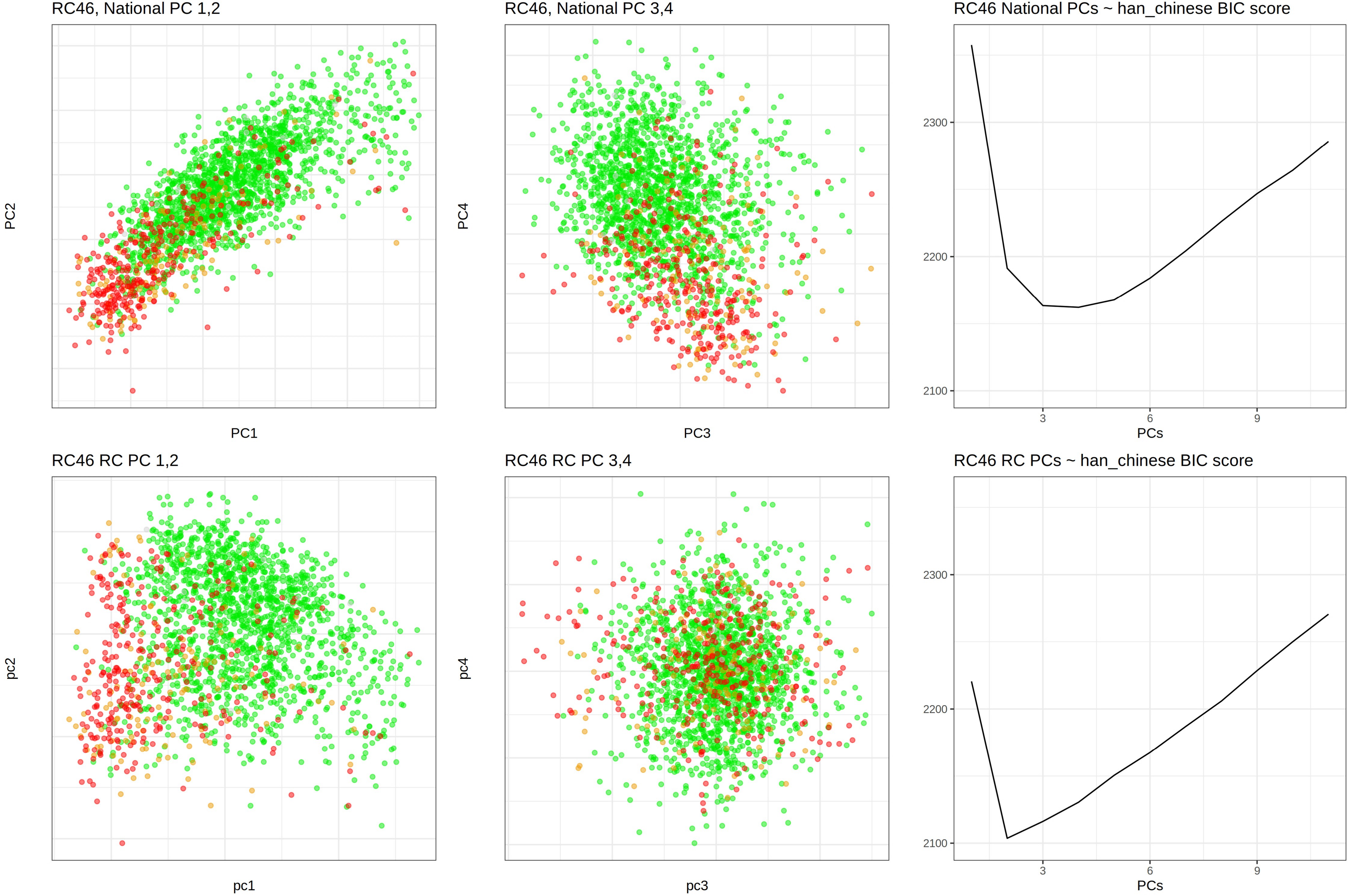

Supplementary Figure S12

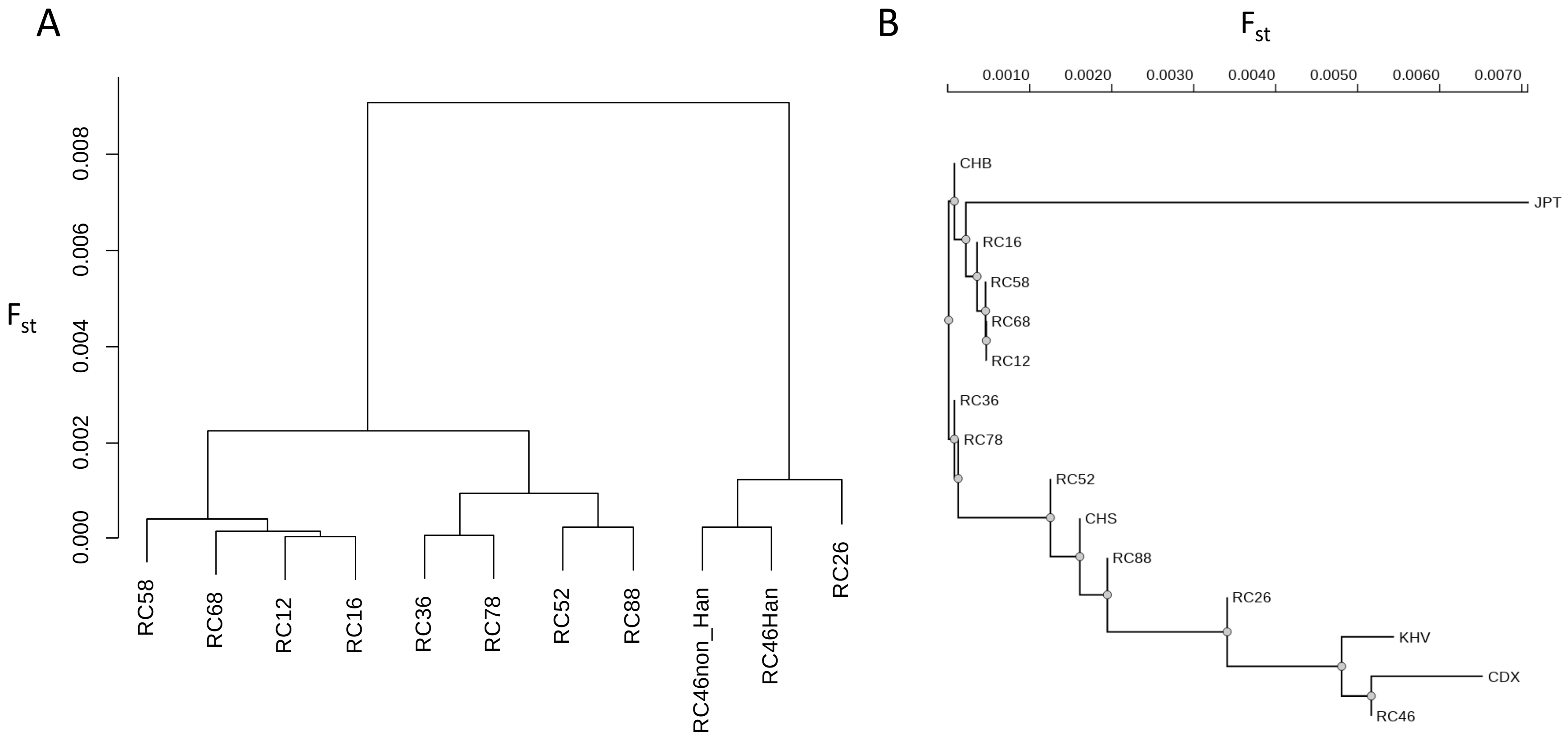

Supplementary Figure S13

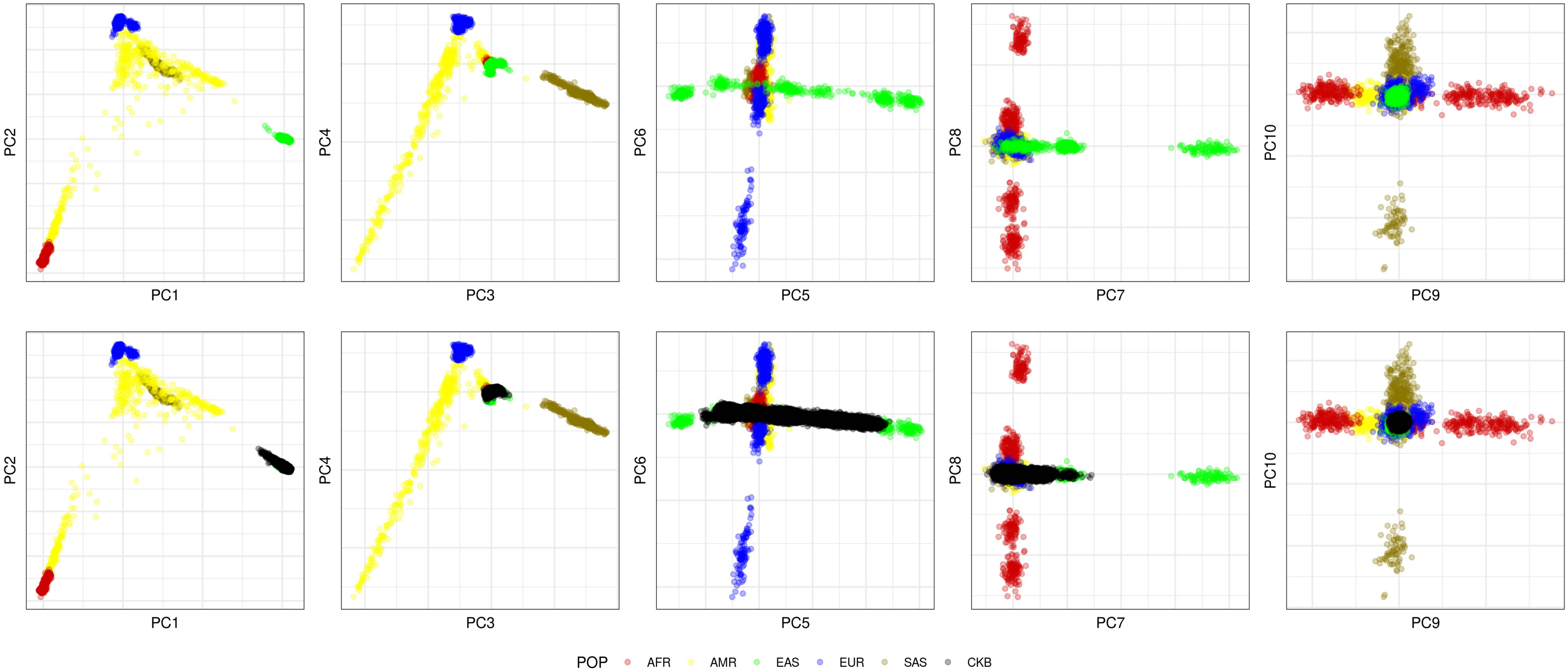

Supplementary Figure S14

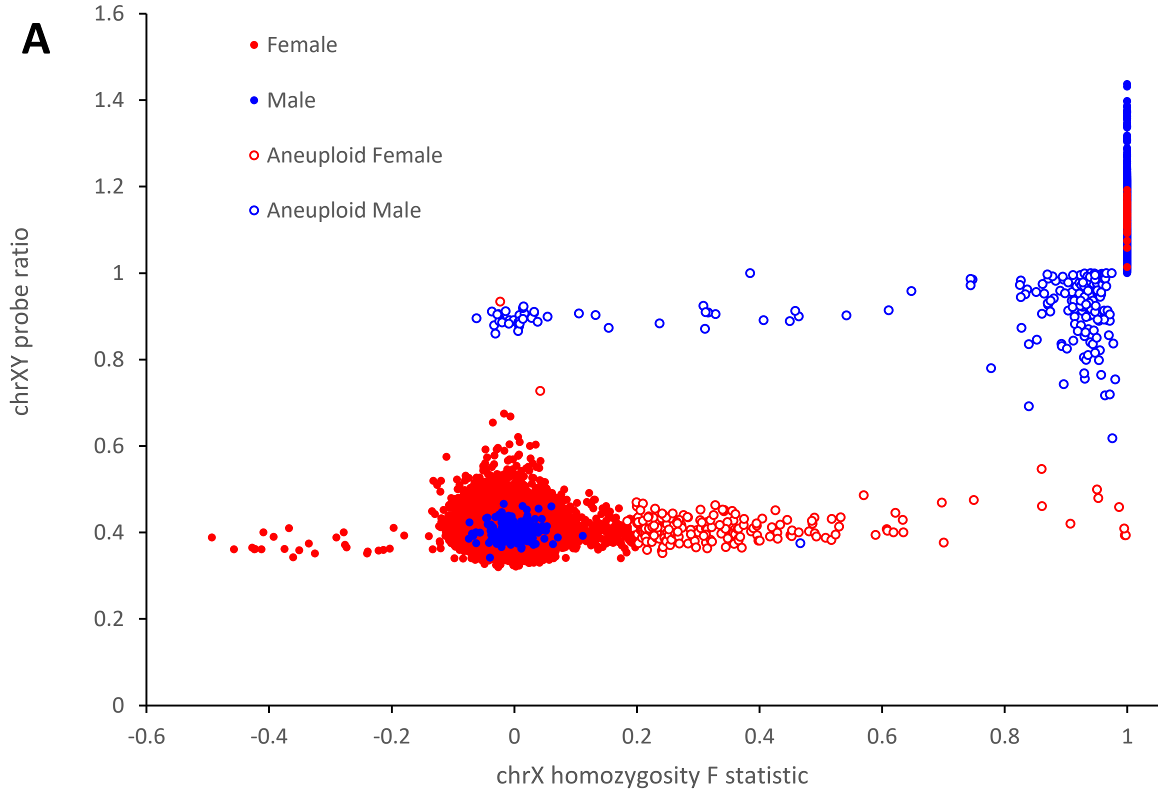


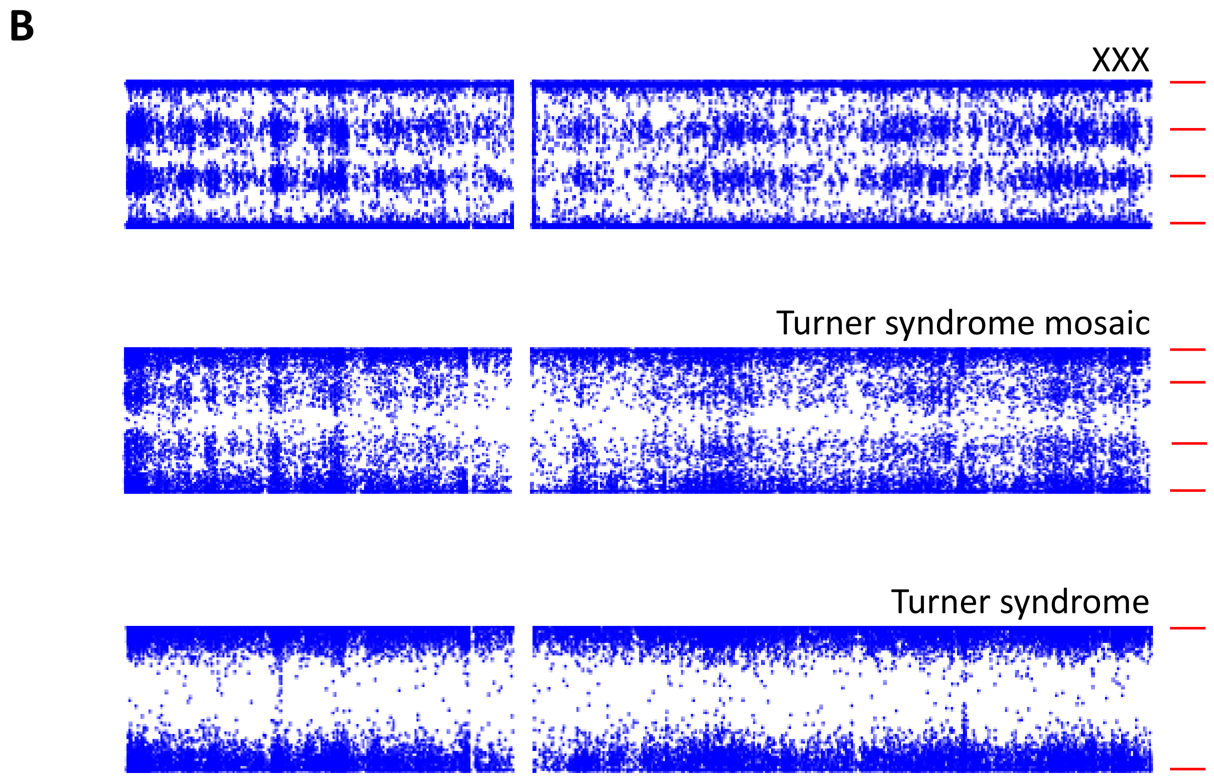

Supplementary Figure S15

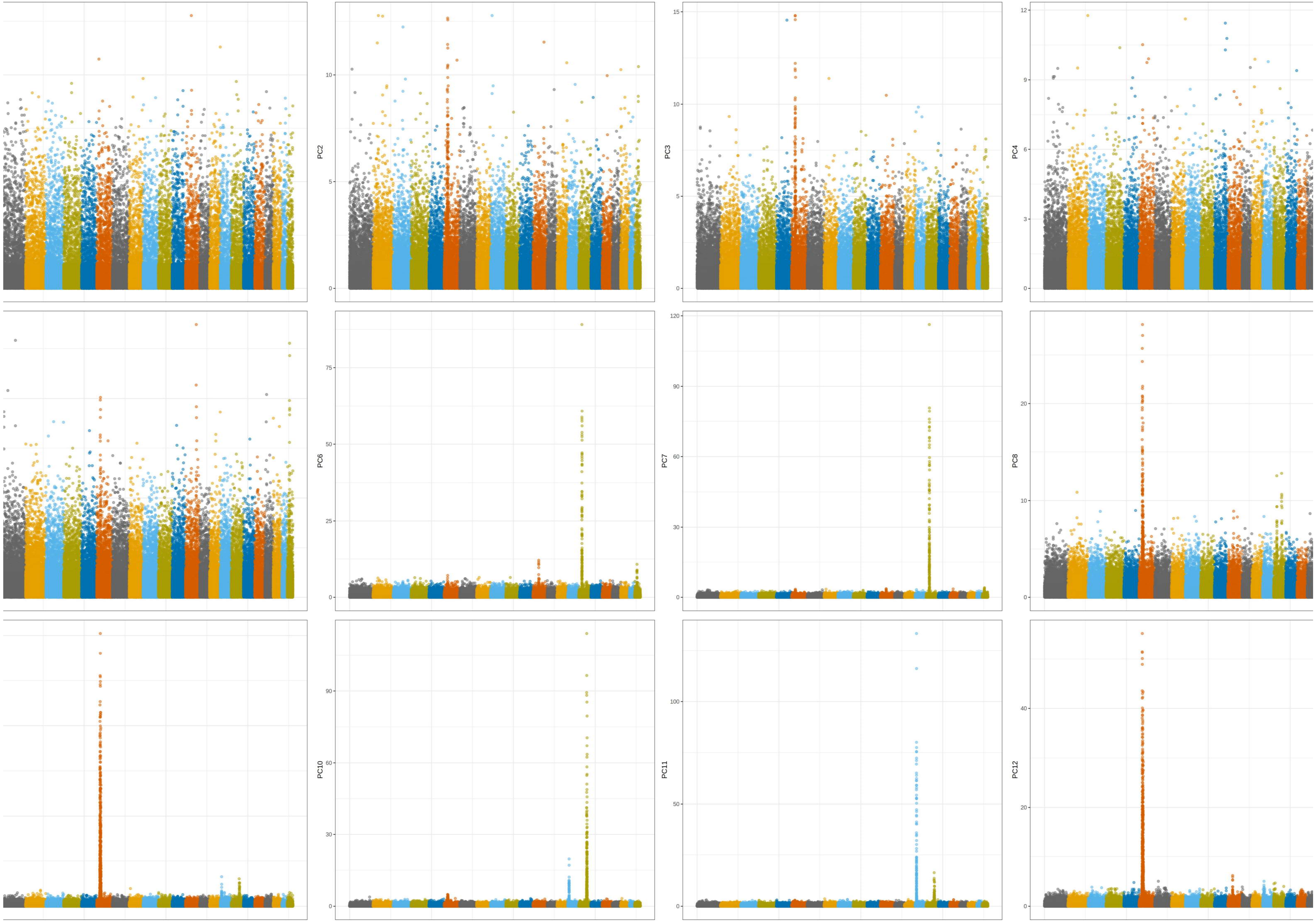

Supplementary Figure S16

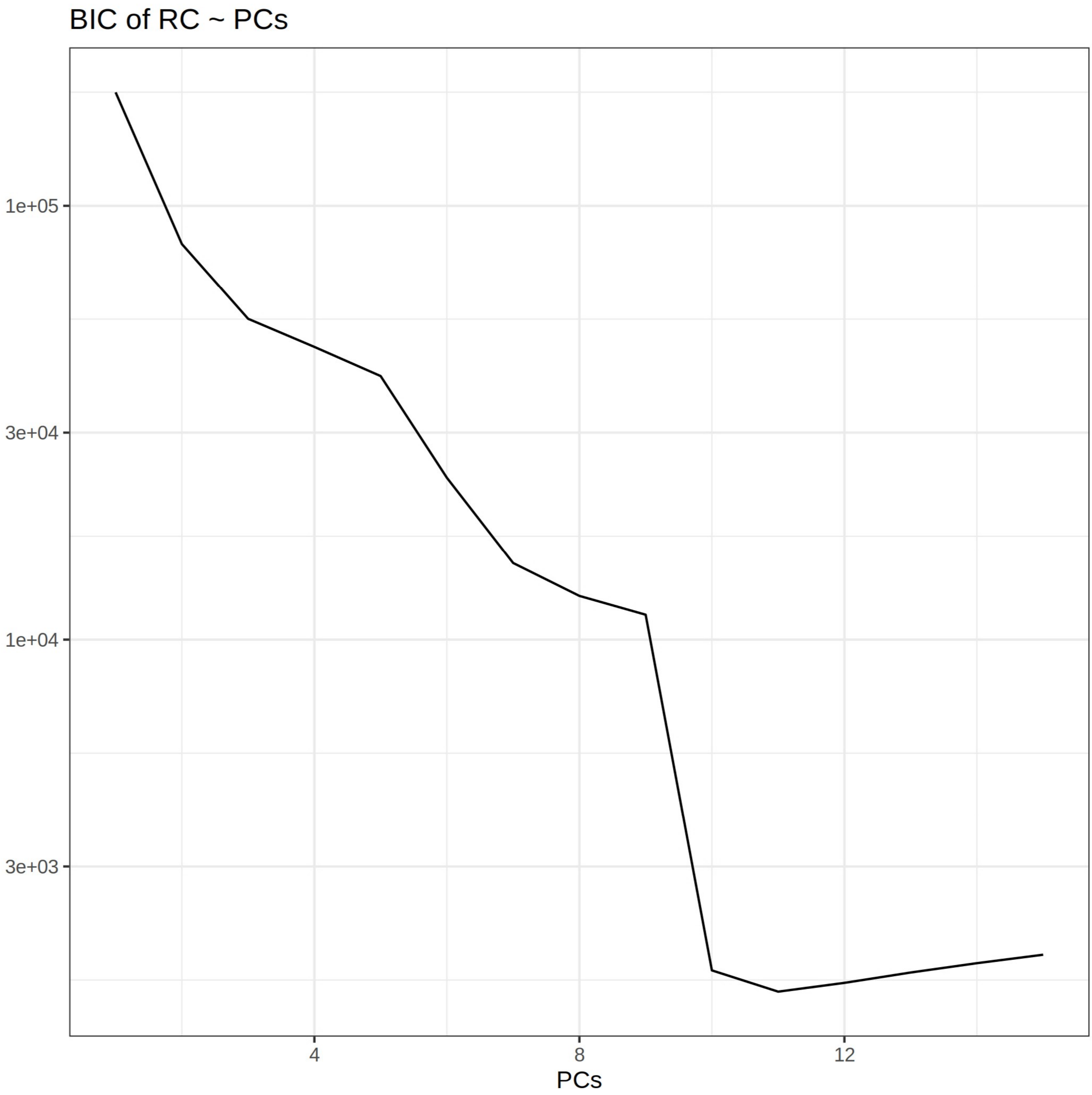
